# Trends in aetiology and antibiotic resistance of bacterial keratitis isolates from South India between 2013-2024

**DOI:** 10.1101/2025.10.01.25337045

**Authors:** Prajna Lalitha, Rameshkumar Gunasekaran, Florence Burté, Namperumalsamy Venkatesh Prajna, Bethany Mills

## Abstract

**Objectives:** Assess bacterial isolate aetiology, antimicrobial (AMR) and multidrug (MDR) resistance trends, cultured from patients with infectious keratitis at a tertiary referral hospital in South India.

**Methods:** This retrospective study examined all-bacterial aetiology, and AMR/MDR profiles of *Streptococcus pneumoniae, Nocardia* spp., *Staphylococcus* aureus and *Pseudomonas aeruginosa* from bacterial keratitis microbiology records (2013-2024). Spearman’s rank correlation was used to identify significant trends over the twelve-year period.

**Results:** *P. aeruginosa* (n=1047) and *S. pneumoniae* (n=987) were the most frequently isolated species. Significant increases in the number of *P. aeruginosa* (r_s_: 0.66; P=0.0219) and *S. aureus* (r_s_: 0.70; P=0.0130) cultured isolates were identified.

Few (significant) AMR trends emerged across the species, and none with first-line bacterial keratitis treatment drugs. However, the number of AMR and MDR *S. aureus* isolates increased significantly (r_s_:0.66; P=0.0235, r_s_:0.63; P=0.0385, respectively) over time. In total, 64% of methicillin-sensitive *S. aureus* (MSSA) and 89% of methicillin-resistant *S. aureus* (MRSA) isolates were resistant to first-line (moxifloxacin) treatment. Conversely, only 2% of *S. pneumoniae* and 1% of *Nocardia* spp. isolates were resistant to first-line treatment. Although *P. aeruginosa* AMR rates increased significantly over time (r_s_:0.83; P=0.0013), 96% of total isolates remained susceptible to first-line treatment.

**Conclusion:** Bacterial aetiology and antibiotic resistance rates changed over time for several key pathogens causing keratitis, most notably for *S. aureus.* Dual therapy (moxifloxacin plus cefotaxime) may better address MSSA infections (98% susceptibility), while the high (57%) MDR MRSA rate may necessitate more tailored treatment plans based on isolate susceptibility reports.

**Summary box:** *What was known before:*

- Antimicrobial resistance poses an increasing threat to the management of microbial keratitis.
- Antimicrobial resistance varies depending on locality, which may influence treatment practices.
- Across India, a high rate of drug-resistant *Pseudomonas aeruginosa* has been reported.

*What this study adds:*

- We report trends in aetiology and antibacterial resistance over 12-years from a tertiary referral clinic in South India.
- The most commonly isolated bacterial species, *Streptococcus pneumoniae*, and *Pseudomonas aeruginosa* remained highly susceptible to most antibiotics. For *P. aeruginosa* this is in contrast with other reports from across India.
- The majority of *Staphylococcus aureus* isolates were resistant to first-line treatment (monotherapy). This was exacerbated for methicillin-resistant strains. Dual-therapy may be advisable for the management of *S. aureus* infections in this region.

## Introduction

Microbial keratitis is an ocular emergency and sight threatening condition affecting over 2 million people each year (1, 2). Microbial keratitis may be caused by a wide variety of opportunistic pathogens; over 680 species of bacteria, fungi, virus and amoeba have been attributed to the disease (3, 4). While this includes 250 bacterial species, the majority of cases are caused by a few species of gram-positive bacteria, primarily *Staphylococcus aureus, Coagulase Negative Staphylococcus* (CoNS) and *Streptococcus pneumoniae*; and gram-negative *Pseudomonas aeruginosa* (5, 6). *Nocardia* spp. is an emerging and important keratitis causing pathogen in some regions of India, where it has been reported as the third most prevalent bacterial species (7, 8).

Broad-spectrum, topical antibiotic therapy remains the gold-standard treatment approach for bacterial keratitis, often in the absence of a microbiological diagnosis. Worldwide, the most common treatment regimens utilise a fluoroquinolone as a monotherapy, or combination dual-therapy of cephalosporin, aminoglycoside and/or fluoroquinolone for refractory infections, as per local guidelines (6, 9, 10). However, antimicrobial resistance (AMR) of keratitis causing bacterial isolates is an increasing public health threat, complicating treatment (5, 6, 11). Correlation between increased *in vitro* antimicrobial minimum inhibitory concentration (MIC) and worse clinical outcomes has been reported (12–15).

The incidence of microbial keratitis, pathogen aetiology and AMR profile are each subject to geographic and temporal variations. It is critical to understand these trends globally and in the local context to inform policy and treatment guidelines. As recently reported by Khor *et al* (as part of the Asia Cornea Society Infectious Keratitis Study (ACSIKS)), and Tuft *et al* (large meta-analysis) *Pseudomonas aeruginosa* isolates from India (or south Asia) were more likely to exhibit AMR compared to samples obtained elsewhere in Asia (16–18). However, their analysis’ combined data from across India/south Asia, irrespective of geographic location. This is despite previous data from South India (collected between 2002-2012) indicating differential resistance patterns of bacterial isolates compared to samples obtained more recently in North, Central and Eastern India (19–23).

Herein, we report aetiology of bacterial keratitis isolates, cultured from patients at a tertiary care centre in South India over a 12-year period (between 2013-2024), as well as report antibiotic resistance trends-over time. We further report total AMR, multidrug resistance (MDR), and susceptibility during this time, in the context of local prescribing practices.

## Materials and Methodology

We conducted a retrospective observational study, utilising microbiological records of patients diagnosed with microbial keratitis at a tertiary eye care referral centre in South India (January 2013 - December 2024). All patients with a positive bacterial culture were included in the study analysis. Microbial aetiology and antimicrobial resistance profiles of culture positive cases were extracted over the 12-year period. Only culture-positive bacterial keratitis cases were included; fungal and mixed infections were excluded. This study was approved by the Aravind Eye Hospital Institutional Review Board, Madurai (RET201300519).

### Microbiological identification of pathogens

Corneal scraping specimens were cultured as per routine clinical practice on solid agar. Following growth, pure cultures of the organisms were used for species level identification through standard microscopic and biochemical tests. Further details are provided in the online supplementary methodology.

### Antibacterial susceptibility testing

Each pure bacterial isolate underwent *in vitro* antibacterial susceptibility testing using the Kirby-Bauer disc diffusion method as per the CLSI guidelines (24). Details of antibacterial agents used are provided in the online supplementary methodology, and zone diameter breakpoints are reported in Supplementary Table S1.

### Statistical analysis

Spearman’s rank correlation coefficient (R_s_) was used to measure the strength and direction of associations over time (see online supplementary methodology for association thresholds). *P_≤_ 0.05; **P_≤_ 0.01; ***P_≤_ 0.001; and ****P_≤_ 0.0001. All analyses were performed using GraphPad Prism version 10 (GraphPad Software, USA). Upset plots were produced using the UpSetR package in R (version R 4.5.0) (25).

## Results

### 1. Trends in bacterial isolation rate and aetiology

A total of 22,887 patient corneal scrapes were cultured over the 12-year period, with a 53.9% culture positivity rate. Fungi were the most frequently cultured organism (8249 isolates, including 358 which also grew bacteria). In total, bacteria grew from 4250 samples (34.5% of culture positive samples). Just 189 samples had parasite growth (2.9% of culture positive samples).

There was no significant trend in the total number of cultured bacterial isolates between 2013-2024 (with a notable reduction in 2020 due to the COVID-19 pandemic). However, over the study period, the number (P=0.0187), and therefore the overall proportion of gram-negative bacteria significantly increased over time (P=0.024) (figure 1a,b, supplementary table S2 and S3).

**Figure 1.**
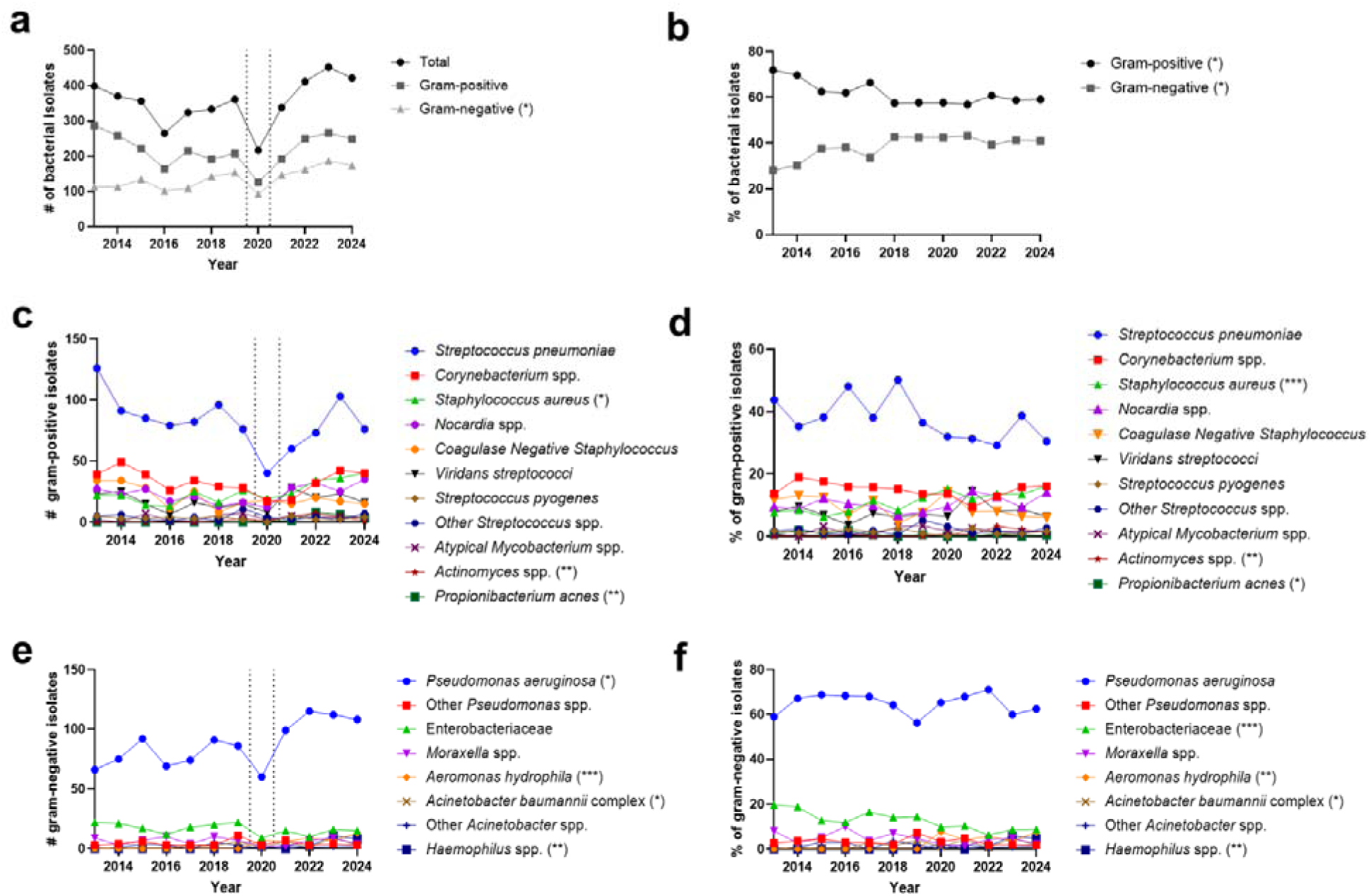
Bacterial isolates cultured from corneal scrape samples 2013-2024. **(a)** Absolute number of isolates; **(b)** proportion of gram-positive and gram-negative isolates. **(c)** Absolute number and **(d)** proportion of gram-positive bacteria cultured from microbial keratitis patients. **(e)** Absolute number and **(f)** proportion of gram-negative bacteria cultured from microbial keratitis patients. Plots display bacteria with a minimum of 20 isolates cultured during the study period. Vertical dashed lines in (a, c and e) indicate where fewer samples were collected due to COVID-19 pandemic reducing patient numbers. Trends analysed by Spearman’s rank correlation. *P _≤_ 0.05; **P _≤_ 0.01; ***P _≤_ 0.001.

#### 1.1. Key gram-positive bacteria trends

A total of 2627 gram-positive bacteria were cultured (supplementary table S4 and S5). *Streptococcus pneumoniae* was consistently isolated with the highest frequency (figure 1c), contributing to an average of 38% of gram-positive cases (figure 1d). *S. aureus* was cultured with increasing frequency over time (P=0.013), doubling from 8% in 2013 to 16% of gram-positive isolates in 2024 (P=0.0005). Notably, *Actinomyces* spp. (P=0.004) and *Propionibacterium acnes* (P=0.0032) significantly increased in number and as a proportion of cultured gram-positive bacteria. While we identified relatively few isolates of *Actinomyces* spp. and 20 *P. acnes*, the majority (18/23 *Actinomyces* spp. and 19/20 *P. acnes* isolates) were cultured between 2020-2024 (supplementary table S4 and S5).

#### 1.2. Key gram-negative bacteria trends

A total of 1623 gram-negative bacteria were cultured (supplementary table S6 and S7), with *Pseudomonas aeruginosa* isolated with the highest frequency (figure 1e). While cases of microbial keratitis caused by *P. aeruginosa* increased significantly over time (P=0.0219), this was proportional to the overall increase in gram-negative organisms cultured from patients, and *P. aeruginosa* contributed steadily to an average of 65% of gram-negative cases throughout the study period (figure 1f).

Conversely, *Enterobacteriaceae* spp. numbers remained relatively stable over the course of the study, and therefore represented a decreasing trend in their proportion of gram-negative bacteria (P=0.0008), halving between 2013 and 2024. Both *Aeromonas hydrophila* (P=0.0003) and *Haemophilus* spp. (P=0.0037) were only cultured from patients from 2020 onwards, contributing to 42 and 20 cases respectively (supplementary table S6 and S7).

### 2. Trends in antimicrobial resistance (AMR) and multidrug resistance (MDR)

#### 2.1. S. pneumoniae

Antibiotic resistance data were available for 960 (97.2%) of the *S. pneumoniae* clinical isolates. The isolates were screened against ten antibiotics from five classes (table 1, supplementary figure S1a, supplementary table S8).

**Table 1.** Trends in bacterial isolate resistance to antibiotics between 2013-2024, analysed by Spearman’s rank correlation.

|  |  |  |  |  |  |  |  |
| --- | --- | --- | --- | --- | --- | --- | --- |
|  |  | **Antibiotic** | **$r_s$** | **95% confidence interval** | **Strength of correlation** | **P** |  |
| **Sp** | Cephalosporins | Cefazolin | 0.64 | 0.09 to 0.89 | **Moderate positive** | **0.0269** | **\*** |
|  |  | Cefotaxime | 0.09 | -0.52 to 0.65 | Negligible | 0.7693 | ns |
|  | Fluoroquinolones | Levofloxacin | 0.15 | -0.48 to 0.68 | Negligible | 0.6459 | ns |
|  |  | Gatifloxacin | 0.15 | -0.48 to 0.68 | Negligible | 0.63 | ns |
|  |  | Moxifloxacin | 0.25 | -0.40 to 0.73 | Weak positive | 0.4392 | ns |
|  |  | Ciprofloxacin | -0.2 | -0.70 to 0.44 | Weak negative | 0.5379 | ns |
|  |  | Ofloxacin | -0.33 | -0.77 to 0.32 | Weak negative | 0.2935 | ns |
|  | Amphenicols | Chloramphenicol | -0.13 | -0.67 to 0.49 | Negligible | 0.6781 | ns |
|  | Tetracyclines | Tetracycline | 0.81 | 0.42 to 0.95 | **Strong positive** | **0.0024** | **\*\*** |
|  | Glycopeptides | Vancomycin | NA |  |  |  |  |
| **No** | Cephalosporins | Cefazolin | 0.61 | 0.04 to 0.88 | **Moderate positive** | **0.0377** | **\*** |
|  |  | Cefotaxime | -0.01 | -0.59 to 0.58 | Negligible | 0.991 | ns |
|  |  | Ceftazidime | -0.01 | -0.60 to 0.58 | Negligible | 0.9689 | ns |
|  | Fluoroquinolones | Levofloxacin | -0.24 | -0.73 to 0.40 | Weak negative | 0.4434 | ns |
|  |  | Gatifloxacin | 0.13 | -0.50 to 0.66 | Negligible | 0.6999 | ns |
|  |  | Moxifloxacin | -0.41 | -0.80 to 0.24 | Moderate negative | 0.1896 | ns |
|  |  | Ciprofloxacin | -0.16 | -0.68 to 0.47 | Negligible | 0.6192 | ns |
|  |  | Ofloxacin | -0.41 | -0.80 to 0.23 | Moderate negative | 0.1845 | ns |
|  | Amphenicols | Chloramphenicol | -0.01 | -0.59 to 0.58 | Negligible | 0.9865 | ns |
|  | Aminoglycosides | Amikacin | -0.24 | -0.72 to 0.41 | Weak negative | 0.4697 | ns |
|  |  | Gentamicin | 0.13 | -0.49 to 0.67 | Negligible | 0.6832 | ns |
|  |  | Tobramycin | 0.38 | -0.26 to 0.79 | Weak positive | 0.2201 | ns |
|  | Glycopeptides | Vancomycin | -0.32 | -0.76 to 0.33 | Weak negative | 0.3155 | ns |
| **Sa** | Cephalosporins | Cefazolin | 0.33 | -0.31 to 0.77 | Weak positive | 0.0888 | ns |
|  |  | Cefotaxime | 0.59 | 0.00 to 0.87 | **Moderate positive** | **0.0489** | **\*** |
|  |  | Cefoxitin | 0.52 | -0.10 to 0.85 | Moderate positive | 0.2886 | ns |
|  | Fluoroquinolones | Levofloxacin | 0.43 | -0.21 to 0.81 | Moderate positive | 0.1616 | ns |
|  |  | Gatifloxacin | 0.48 | -0.15 to 0.83 | Moderate positive | 0.1154 | ns |
|  |  | Moxifloxacin | 0.36 | -0.29 to 0.78 | Weak positive | 0.256 | ns |

|  |  |  |  |  |  |  |  |
| --- | --- | --- | --- | --- | --- | --- | --- |
|  |  | Ciprofloxacin | 0.34 | -0.31 to 0.77 | Weak positive | 0.2869 | ns |
|  |  | Ofloxacin | 0.38 | -0.26 to 0.79 | Weak positive | 0.2183 | ns |
|  | Amphenicols | Chloramphenicol | 0.37 | -0.27 to 0.79 | Weak positive | 0.233 | ns |
|  | Aminoglycosides | Gentamicin | -0.19 | -0.70 to 0.44 | Negligible | 0.5443 | ns |
|  |  | Tobramycin | -0.15 | -0.68 to 0.48 | Negligible | 0.6389 | ns |
|  | Glycopeptides | Vancomycin | -0.30 | -0.75 to 0.35 | Weak negative | 0.3423 | ns |
| MSSA | Cephalosporins | Cefazolin | 0.24 | -0.40 to 0.72 | Weak positive | 0.4606 | ns |
|  |  | Cefotaxime | 0.03 | -0.57 to 0.60 | Negligible | 0.9414 | ns |
|  | Fluoroquinolones | Levofloxacin | 0.22 | -0.42 to 0.71 | Weak positive | 0.499 | ns |
|  |  | Gatifloxacin | 0.45 | -0.19 to 0.82 | Moderate positive | 0.144 | ns |
|  |  | Moxifloxacin | 0.20 | -0.44 to 0.70 | Weak positive | 0.5397 | ns |
|  |  | Ciprofloxacin | 0.13 | -0.50 to 0.66 | Negligible | 0.6999 | ns |
|  |  | Ofloxacin | 0.10 | -0.52 to 0.65 | Negligible | 0.7613 | ns |
|  | Amphenicols | Chloramphenicol | 0.45 | -0.19 to 0.82 | Moderate positive | 0.1451 | ns |
|  | Aminoglycosides | Gentamicin | 0.12 | -0.50 to 0.66 | Negligible | 0.7147 | ns |
|  |  | Tobramycin | -0.03 | -0.60 to 0.57 | Negligible | 0.9388 | ns |
|  | Glycopeptides | Vancomycin | -0.37 | -0.79 to 0.28 | Weak negative | 0.2576 | ns |
| MRSA | Cephalosporins | Cefazolin | 0.13 | -0.52 to 0.69 | Negligible | 0.7009 | ns |
|  |  | Cefotaxime | -0.22 | -0.74 to 0.45 | Weak negative | 0.5027 | ns |
|  | Fluoroquinolones | Levofloxacin | 0.20 | -0.47 to 0.72 | Weak positive | 0.5611 | ns |
|  |  | Gatifloxacin | -0.22 | -0.73 to 0.45 | Weak negative | 0.517 | ns |
|  |  | Moxifloxacin | -0.18 | -0.71 to 0.49 | Negligible | 0.5945 | ns |
|  |  | Ciprofloxacin | -0.46 | -0.84 to 0.22 | Moderate negative | 0.1596 | ns |
|  |  | Ofloxacin | -0.46 | -0.84 to 0.22 | Moderate negative | 0.1596 | ns |
|  | Amphenicols | Chloramphenicol | -0.05 | -0.64 to 0.58 | Negligible | 0.8978 | ns |
|  | Aminoglycosides | Gentamicin | -0.90 | -0.98 to -0.65 | Very strong negative | 0.0003 | *** |
|  |  | Tobramycin | -0.81 | -0.95 to -0.40 | Strong negative | 0.0035 | ** |
|  | Glycopeptides | Vancomycin | 0.03 | -0.60 to 0.63 | Negligible | 0.9636 | ns |
| Pa | Cephalosporins | Cefotaxime | 0.6 | 0.023 to 0.88 | Moderate positive | 0.0428 | * |
|  |  | Ceftazidime | -0.9 | -0.97 to -0.67 | Very strong negative | 0.0002 | *** |
|  | Fluoroquinolones | Levofloxacin | 0.24 | -0.41 to 0.72 | Weak positive | 0.4573 | ns |
|  |  | Gatifloxacin | -0.45 | -0.82 to 0.18 | Moderate negative | 0.1404 | ns |
|  |  | Moxifloxacin | -0.19 | -0.70 to 0.45 | Negligible | 0.5577 | ns |
|  |  | Ciprofloxacin | -0.22 | -0.72 to 0.42 | Weak negative | 0.4851 | ns |
|  |  | Ofloxacin | -0.64 | -0.89 to -0.09 | Moderate negative | 0.0278 | * |
|  | Aminoglycosides | Amikacin | -0.77 | -0.93 to -0.33 | Strong negative | 0.0049 | ** |

|  |  |  |  |  |  |  |  |
| --- | --- | --- | --- | --- | --- | --- | --- |
|  |  | Gentamicin | -0.65 | -0.90 to -0.10 | **Moderate negative** | **0.0257** | **\*** |
|  |  | Tobramycin | -0.67 | -0.90 to -0.14 | **Moderate negative** | **0.0205** | **\*** |
|  | Polymyxins | Polymyxin B | -0.32 | -0.76 to 0.33 | Weak negative | 0.3104 | ns |

Sp: *S. pneumoniae*; No: *Nocardia* spp.; Sa: *S. aureus*; MSSA: Methicillin sensitive *S. aureus*; MRSA: Methicillin Resistant *S. aureus*; Pa: *P. aeruginosa*. Significant ( $P < 0.05$ ) trends depicted in bold.

Resistance towards tetracycline was the most commonly found resistance for *S. pneumoniae* in 2013, observed in 32% of isolates. Over the study period, there was a strong upward trend in resistance rates (P=0.0024), peaking at 58% in 2022.

While resistance towards the fluoroquinolones was less frequently observed, there was a peak in resistance towards ciprofloxacin, gatifloxacin and ofloxacin between 2017-2019, where over one third of isolates were resistant. The proportion of isolates resistant to these drugs dropped to pre-2016 levels from 2020 and have remained stable. Levofloxacin and moxifloxacin resistance was consistently observed in 10% or fewer isolates across the study period.

*S. pneumoniae* remained highly susceptible to cefotaxime throughout the study period, with 99% of isolates showing susceptibility. While 99% of isolates were also susceptible to cefazolin over the study period, there has been a recent notable, moderate-increase in the proportion of resistant isolates (P=0.0269). Resistance towards chloramphenicol remained low and stable, with an average of 96% of isolates showing susceptibility. None of the isolates demonstrated resistance to vancomycin.

Between 2013-2024, there was no significant change in the number or overall proportion of *S. pneumoniae* isolates displaying AMR (resistance to 1-2 classes of antibiotic) or MDR (resistance to _≥_3 antibiotic classes) (figure 2a). Across the study, 37% of the *S. pneumoniae* isolates were susceptible to all of the drugs tested, 45% were resistant to a single class, 17% to two classes and 2% were resistant to 3 or more classes, making them MDR (figure 3a).

**Figure 2.**
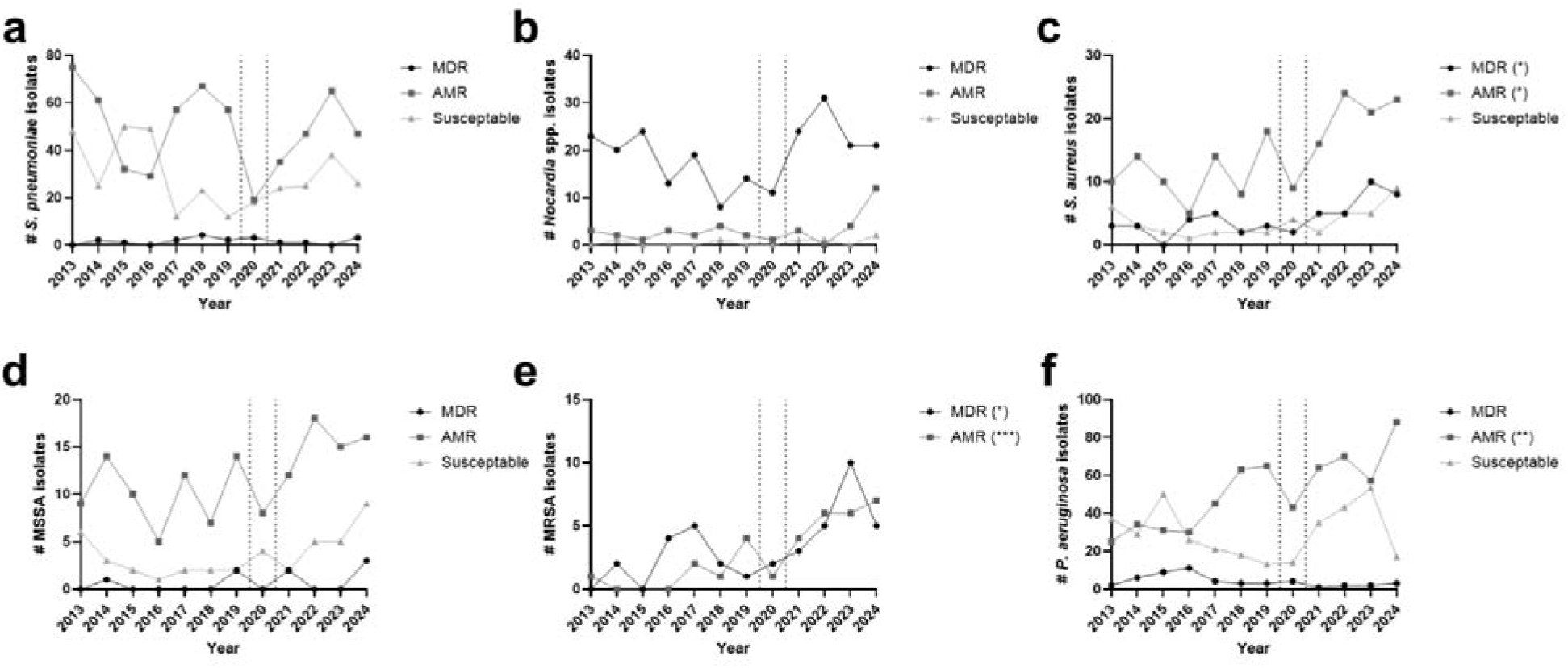
Bacterial isolates cultured from corneal scrape samples 2013-2024 categorised by antimicrobial resistance status. Data are presented as the number of (a) *S. pneumoniae*; **(b)** *Nocardia* spp; **(c)** *S. aureus* (all); **(d)** methicillin sensitive *S. aureus* (MSSA); **(e)** methicillin resistant *S. aureus* (MRSA); and **(f)** *P. aeruginosa* isolates that were susceptible to all drugs evaluated, those with resistance to 1-2 classes of antibiotic (AMR), and those with resistance to _≥_3 classes of antibiotic (MDR). Trends analysed by Spearman’s rank correlation. *P _≤_ 0.05; **P _≤_ 0.01; *** P _≤_ 0.001.

**Figure 3.**
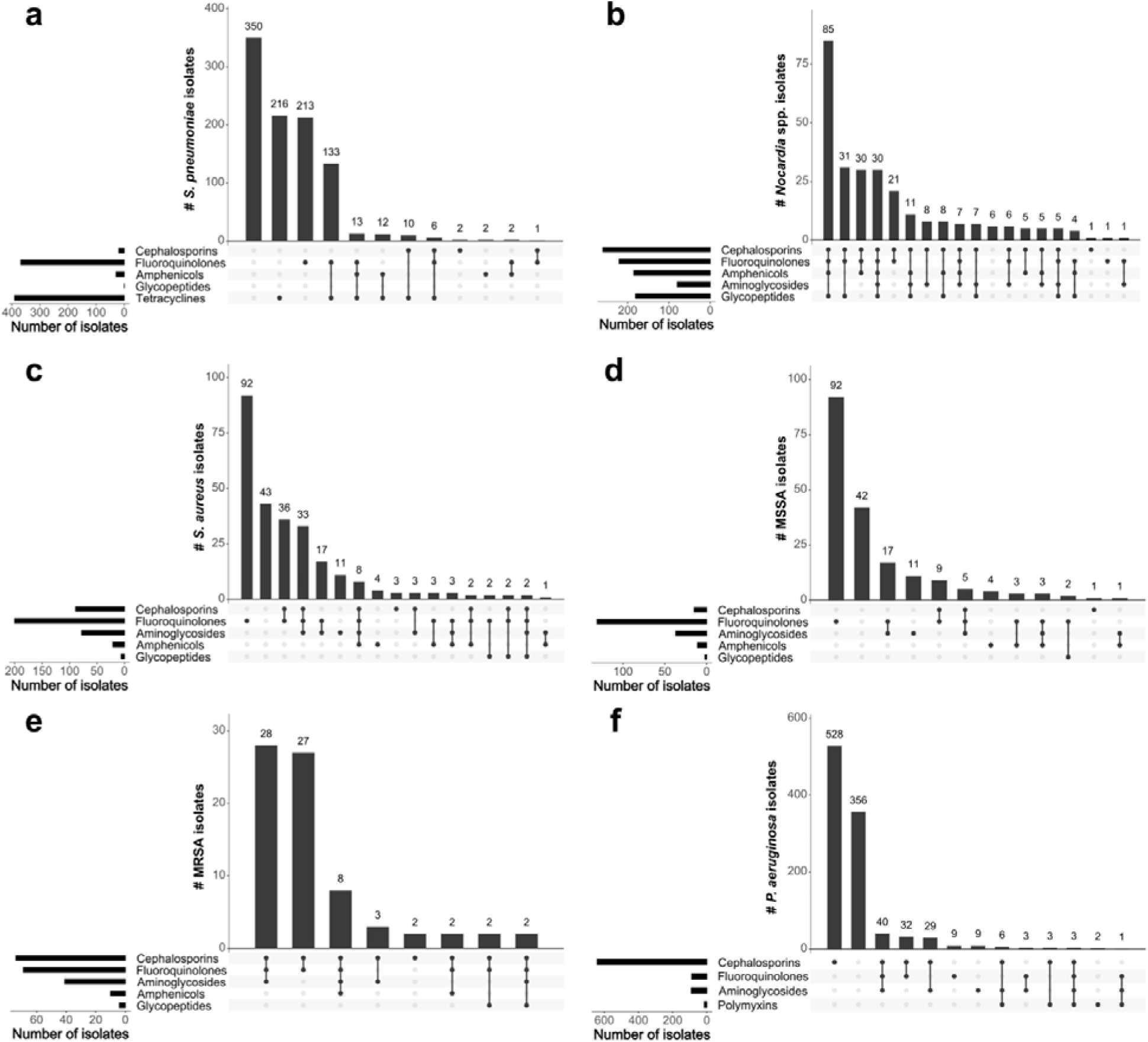
Concurrent resistance of bacterial isolates to antibiotic classes. Upset plots depicting the number of **(a)** *S. pneumoniae*; **(b)** *Nocardia* spp; **(c)** *S. aureus* (all); **(d)** methicillin sensitive *S. aureus* (MSSA); **(e)** methicillin resistant *S. aureus* (MRSA); and **(f)** *P. aeruginosa* isolates with resistance to one or more antibiotic from each class of antimicrobials.

#### 2.2. Nocardia spp

Antibiotic resistance data were available for 272 (98.6%) of the *Nocardia* spp. isolates. Thirteen antibiotics from five classes were evaluated (table 1, supplementary figure S1b, supplementary table S9).

High levels of resistance prevailed for the *Nocardia* isolates. Notably, the *Nocardia* isolates showed a largely erratic resistance pattern to the majority of antimicrobials tested, and there were no significant changes over time, with the exception of cefazolin. For this antibiotic, there was a moderate, significant increase in the proportion of resistant isolates over time (P=0.0377). Overall, aminoglycosides remained the most effective class of antibiotics against *Nocardia* spp., with ∼75% of isolates susceptible to gentamicin and tobramycin, and 99% to amikacin.

There was no significant trend in AMR or MDR *Nocardia* spp. isolates between 2013-2024 (figure 2b). Across the study, just 2% of the *Nocardia* isolates were found to be susceptible to all of the drugs tested, <1% were resistant to a single class, 13% to two classes and 84% were MDR.

#### 2.3. S. aureus

Antibiotic resistance data were available for 264 *S. aureus* isolates (91.0%). In total, twelve antibiotics from five classes were screened. Methicillin susceptibility was determined via assay with cefoxitin (26) (table 1, supplementary figure S1c-e, supplementary table S4 and S5). In total, 190 (72%) isolates were methicillin sensitive (MSSA), and 74 (28%) isolates were methicillin resistant (MRSA). We found that while the number MRSA isolates did significantly increase (P=0.0053) over the study period, this was not significant as a proportion of all *S. aureus* isolates due to concurrent increasing (non-significant) incidence of MSSA.

Across all *S. aureus* isolates, cefotaxime was the only drug for which the proportion of resistant isolates significantly increased over time (P=0.0489, table 1, supplemental figure S1c, supplemental table S10). 72% of isolates resistant to one or more fluoroquinolone. Across all *S. aureus* isolates, there was a moderate increase in the number of AMR and MDR isolates over time (P<0.05, figure 2c).

There were no significant trends in resistance rates MSSA to any tested antibiotic over the study period (table 1, figure 2d, supplemental figure S1d, supplemental table S11), whereas there was a significant reduction in the proportion of aminoglycoside-resistant MRSA isolates (table 1, supplemental figure S1e, supplemental table S12). Despite this, the total number of AMR and MDR MRSA increased over time (figure 2e). However, MRSA trends should be interpreted with caution due to low isolate numbers, with just 74 reported over the 12-year study period (table 1). Across all MSSA isolates, 22% were susceptible to all antimicrobials tested, 57% were resistant to one class, 17% were resistant to two antimicrobial classes, and 4% were MDR. In contrast, 57% of MRSA isolates were MDR, and 41% were resistant to two classes of antimicrobial (figure 3d,e).

#### 2.4. P. aeruginosa

Antibiotic resistance data were available for 1021 (97.5%) of the *P. aeruginosa* isolates. The isolates were evaluated against eleven antibiotics, from four classes (table 1, supplementary figure S1f, supplementary table S13).

Overall, *P. aeruginosa* remained susceptible to the majority of antibiotics tested, with >90% of isolates susceptible to aminoglycosides; fluoroquinolones; polymyxin B, and ceftazidime. While susceptibility remained high for these antibiotics, decreasing resistance trends of: moderate (ofloxacin (0.0278), gentamycin (P=0.0257), tobramycin (P=0.0205)); strong (amikacin (P=0.0049)) and very strong (ceftazidime (P=0.0002)) strength were observed. While resistance towards ceftazidime was low, high rates of *P. aeruginosa* resistance towards cefotaxime was measured, which moderately increased over time (P=0.0428), leading to a significant increase in the number of AMR *P. aeruginosa* (P=0.0013) (figure 2f). Overall, 54% of *P. aeruginosa* isolates were resistant to one class of antibiotic, 7% were resistant to two classes, and 5% were MDR. 35% of isolates remained susceptible to all antibiotics tested (figure 3f).

#### 2.5. Implications for patient treatment

While AMR susceptibility testing with a broad panel of antibiotics is necessary to characterise each isolate, the treatment options for bacterial keratitis are limited. Therefore, we explored susceptibility and resistance of our isolates in the context of likely prescribing.

In our clinic, *S. pneumoniae* and *S. aureus* are first empirically treated with moxifloxacin. For refractory cases, a cephalosporin (e.g. cefotaxime) may be prescribed in combination with moxifloxacin, or chloramphenicol may be used in combination with polymyxin B.

93% of the *S. pneumoniae* isolates were susceptible to all of these clinically relevant drugs. 98% of the isolates were susceptible to first-line moxifloxacin, and none were resistant to the combination of moxifloxacin and cefotaxime (figure 4a).

**Figure 4.**
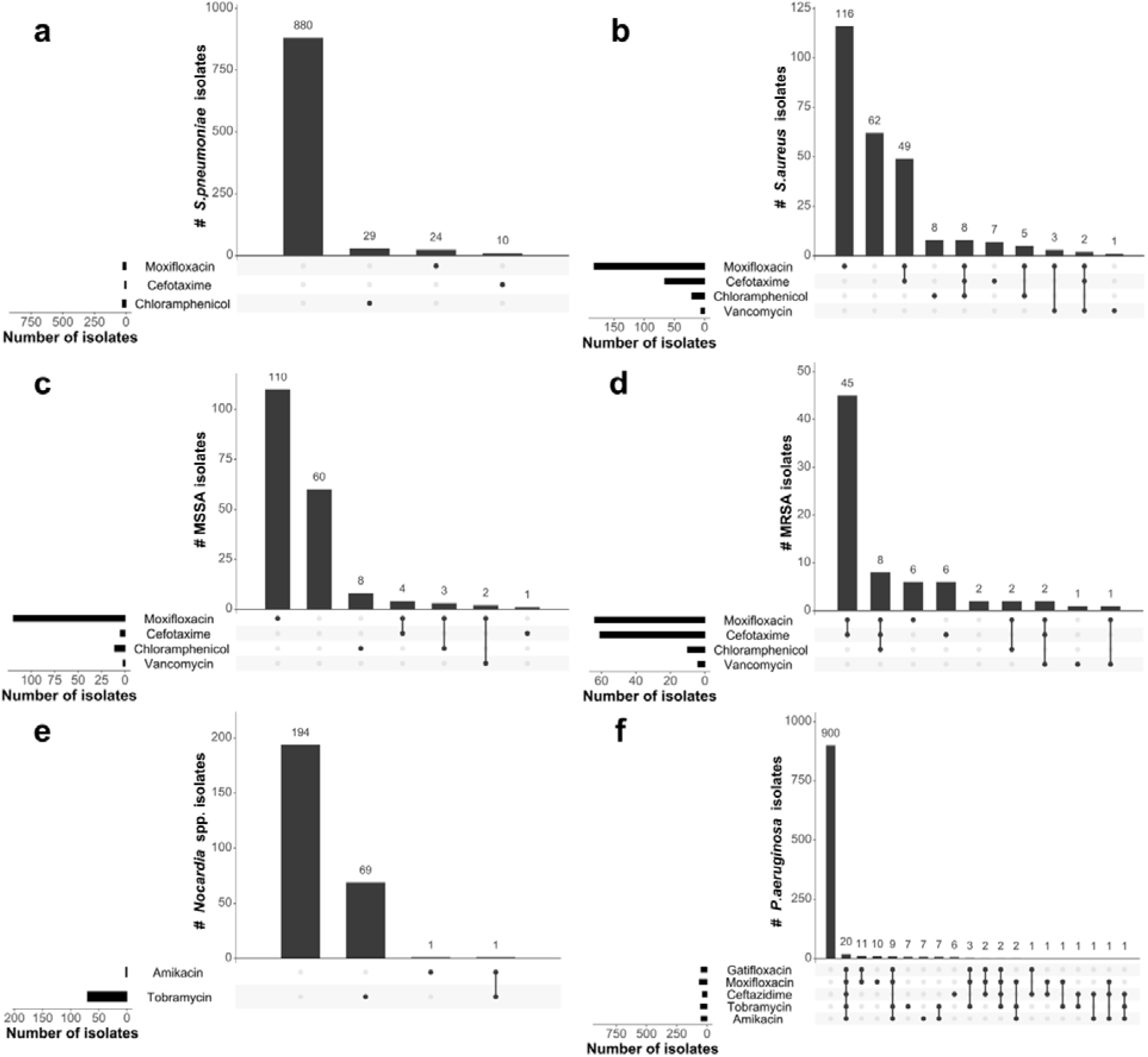
Concurrent resistance of bacterial isolates to clinically-used antibiotics. Upset plots depicting the number of **(a)** *S. pneumoniae*; **(b)** *S. aureus* (all); **(c)** methicillin sensitive *S. aureus* (MSSA); **(d)** methicillin resistant *S. aureus* (MRSA); **(e)** *Nocardia* spp; and **(f)** *P. aeruginosa* isolates with resistance to one or more antibiotic used in clinical management of bacterial keratitis.

Overall, 71% of the *S. aureus* isolates were resistant to first-line moxifloxacin (figure 4b). However, the susceptibility landscape for MSSA and MRSA were divergent. For MSSA, 64% of the isolates were resistant to moxifloxacin, but only 2% of the isolates were resistant to moxifloxacin and cefotaxime. None of the isolates were resistant to all of the standard therapeutic drugs, and 32% of the isolates were susceptible to all of them (figure 4c).

In contrast to the MSSA strains, 88% of the MRSA isolates were resistant to moxifloxacin, and 75% were resistant to the combination of moxifloxacin plus cefotaxime. Just three isolates (4%) were sensitive to all of the clinically applicable antimicrobials, and 11% were resistant to all of them. However, each of these isolates was susceptible to vancomycin, which is reserved as a last-line antimicrobial for MRSA strains. There were four vancomycin-resistant MRSA strains. Three of these were resistant to moxifloxacin, and two were further resistant to cefotaxime. All four remained susceptible to chloramphenicol (figure 4d).

The first line treatment for *Nocardia* spp. keratitis is amikacin, with tobramycin used as a secondary or alternative topical treatment. In our study, 99% of the *Nocardia* spp. isolates were susceptible to amikacin, with 73% of the isolates susceptible to both amikacin and tobramycin. Only one isolate was resistant to both drugs (figure 4e).

In our clinic, keratitis caused by gram-negative bacteria (*P. aeruginosa*) are initially treated with tobramycin and gatifloxacin (or moxifloxacin). These may later be modified to amikacin and ceftazidime. Overall, 96% of the isolates were susceptible to the first-line drug treatment combination. This includes 90.6% of the isolates which were sensitive to all of the clinically relevant drugs. 95% of the isolates were susceptible to gatifloxacin as monotherapy, and 2% of the isolates were resistant to all of these antibiotics (figure 4f).

## Discussion

Microbial keratitis prevalence and treatment remains a significant challenge. While we determined that the five most frequently cultured bacteria were *P. aeruginosa* (25%); *S. pneumoniae* (23%); *Corynebacterium* spp. (9%); *S. aureus* (7%) and *Nocardia* spp. (6%) from our South Indian cohort, a similarly sized study from North India found coagulase-negative *S. aureus* to be the most prevalent organism (61%), followed by *P. aeruginosa* (14%), *S. aureus* (14%), *Enterobacteriaceae* (9%) and *Streptococcus* spp. (2%) (20). This differential aetiology was also observed in Central India, where *S. pneumoniae* was the most prevalent bacteria (43%), followed by *S. aureus* (20%), *Bacillus* spp. (15%) and *P. aeruginosa* (12%) (27). Notably, a 30 year study from another large centre in South India found that their *S. pneumoniae* cases increased over time, and was the only bacteria to show a significant trend between 1991 and 2020 (28), in contrast, our hospital did not see a significant increase in *S. pneumoniae* culture rates over our 12-year study period. We did, however, observe a significant increase in the incidence of culture-positive gram-negative keratitis over the study period, with a strong correlation between *P. aeruginosa* isolate numbers and time.

Although we did see an increase in the absolute numbers of MRSA isolates over time, the number of MSSA isolates also increased. Reassuringly, we did not see an increase in the proportion of MRSA over the study period (after a significant increase between 2002-2012 (29)). Between 2013-2024, 28% of *S. aureus* isolates demonstrated methicillin resistance, which is within the range of other studies from Asia (30, 31), and the USA (32), who have also reported a stabilisation of MRSA isolates. In addition to differential aetiology, varying antibiotic susceptibility profiles were reported across each of these regions, underscoring the need to understand the specific geographical, local and temporal context for microbial keratitis diagnosis and treatment strategy.

Overall, our study identified increasing trends in antimicrobial resistance rates for only a few of the evaluated antibiotics, the majority of which were not first-line treatment choices for treating microbial keratitis. Only *S. aureus* isolates showed a moderate, significant increase in resistance to the cephalosporin, cefotaxime, over-time. However, this trend was lost when MSSA and MRSA were considered separately (likely due to sample size). Somewhat unexpectedly, decreasing trends in resistance were identified for *P. aeruginosa* isolates across several antibiotic classes.

As has been identified by us and others (21, 30, 33), there was a high proportion of fluoroquinolone resistant by *S. aureus* keratitis isolates, even at the outset of this study. *S. aureus* fluoroquinolone resistance is correlated with increasing incidence of multi-drug-resistant MRSA strains (30, 32), and we found that to be true here. 84% of the MRSA isolates were resistant to moxifloxacin, compared to 64% of the MSSA strains.

Increasing fluoroquinolone resistance is particularly concerning for the management of patients, with 0.5% moxifloxacin monotherapy being the first-line treatment for bacterial corneal ulcers (excluding *Nocardia*) in many places. Moreover, clinical isolates from patients who had been treated with fluoroquinolones prior to having their sample collected have been reported to exhibit higher minimum inhibitory concentrations (MICs), compared to clinical isolates obtained from treatment naïve patients (34), indicating that increasing resistance could be driven by the therapy itself. Furthermore, higher *in vitro* moxifloxacin MIC values have been associated with worse clinical outcomes for bacterial keratitis patients (12, 35).

*S. pneumoniae* and *P. aeruginosa* were the most frequently cultured bacteria within our study; accounting for almost half of bacterial keratitis isolates. In contrast to *S. aureus* and *Nocardia* spp., these isolates exhibited higher levels of *in vitro* susceptibility to most of the antibiotics tested. While others have reported increasing antibiotic-resistant *P. aeruginosa* from microbial keratitis patients in India in recent years, particularly in northern and central regions (17, 20–22), we did not find this trend to be true in our South Indian population. Our AMR data reflects that previously reported within our clinic 2002-2013 (23), with >90% susceptibility to each antibiotic, except for cefotaxime observed. Surprisingly, resistance rates towards ceftazidime, ofloxacin, amikacin, gentamicin and tobramycin significantly declined over the last 12 years. While others in more northerly regions of South India also reported 30% resistance rates of *P. aeruginosa* isolates to ceftazidime (28), our observations reflect those found in other regions across Asia (17, 18, 31), and further underscore the need for assessing AMR in the local context.

As described, antibiotic monotherapy is the most common approach for treating bacterial keratitis caused by gram-positive bacteria (10). This approach is supported by our data for *Nocardia* spp., and *S. pneumoniae,* however the landscape may be more complicated for *S. aureus*. Our data indicates that the majority of isolates were resistant to moxifloxacin monotherapy, yet just 2% of MSSA isolates were resistant to moxifloxacin and cefotaxime, therefore dual therapy may be indicated to treat these infections. In contrast 57% of the MRSA isolates were MDR, and may necessitate more tailored treatment plans based on individual isolate susceptibility reports.

However, these decisions should be further informed by clinical response to prescribed treatment regimens for keratitis. This is due to inherent limitations in interpreting antimicrobial resistance data in a clinically meaningful way due to the paucity of CLSI or EUCAST MIC breakpoint guidelines for ocular microbiology. Antibiotics for treating bacterial keratitis are applied topically and frequently, but also have to contend with limited cornea penetration and immediate dilution from tears, therefore local tissue concentrations likely differ from serum concentrations for which the guideline thresholds were derived (36, 37). Thus, we (and others) may have over- or under-reported clinically relevant antibiotic-resistant rates. Nonetheless, the breakpoints that we have utilised are common with other studies reporting on microbial keratitis isolates. Despite this, work to determine standardised, clinically relevant breakpoints are warranted to facilitate rational clinical guidelines for bacterial keratitis.

Nonetheless, our study has many strengths, including the large sample size, 12-year study period and reporting of concurrent resistance profiles across each clinical isolate. We have reported resistance data across a broad panel of antibiotics from multiple classes, beyond those used routinely to treat bacterial keratitis. In a landscape of fluctuating antimicrobial resistance, understanding how stable resistance patterns are (or are not), is essential to identify emerging pathogens and resistance threats. These data are important to closely monitor to guide treatment decisions not only for bacterial keratitis, but other bacterial infections present within the same communities.

Finally, while it is encouraging that antimicrobial susceptibility of the most common bacterial species remains high to first-line treatments, it is of concern that the majority of *S. aureus* isolates from our study were fluroquinolone resistant. Moxifloxacin resistance is associated with worse outcomes (35). Our data supports that dual-therapy may be beneficial for treating *S. aureus* infections in this geographic region.

## Author Contributions (CRediT)

PL: Conceptualization, Data curation, Methodology, Supervision, Writing – review and editing. RG: Data curation, Investigation, Writing – review and editing. FB: Formal analysis, Data curation; Writing – review and editing. NVP: Conceptualization, Data curation, Methodology, Supervision, Writing – review and editing. BM: Conceptualization; Funding acquisition, Formal analysis, Data curation, Methodology, Supervision, Writing – original draft.

## Data availability statement

Data are available on reasonable request. Supplementary information is available at Eye’s website.

For the purpose of open access, the author has applied a Creative Commons Attribution (CC BY) licence to any Author Accepted Manuscript version arising from this submission.

## Data Availability

Data are available on reasonable request to the corresponding authors.

