## Supplemental tables and figures for "Trends in aetiology and antibiotic resistance of bacterial keratitis isolates from South India between 2013-2024"

### Supplementary Figure 1

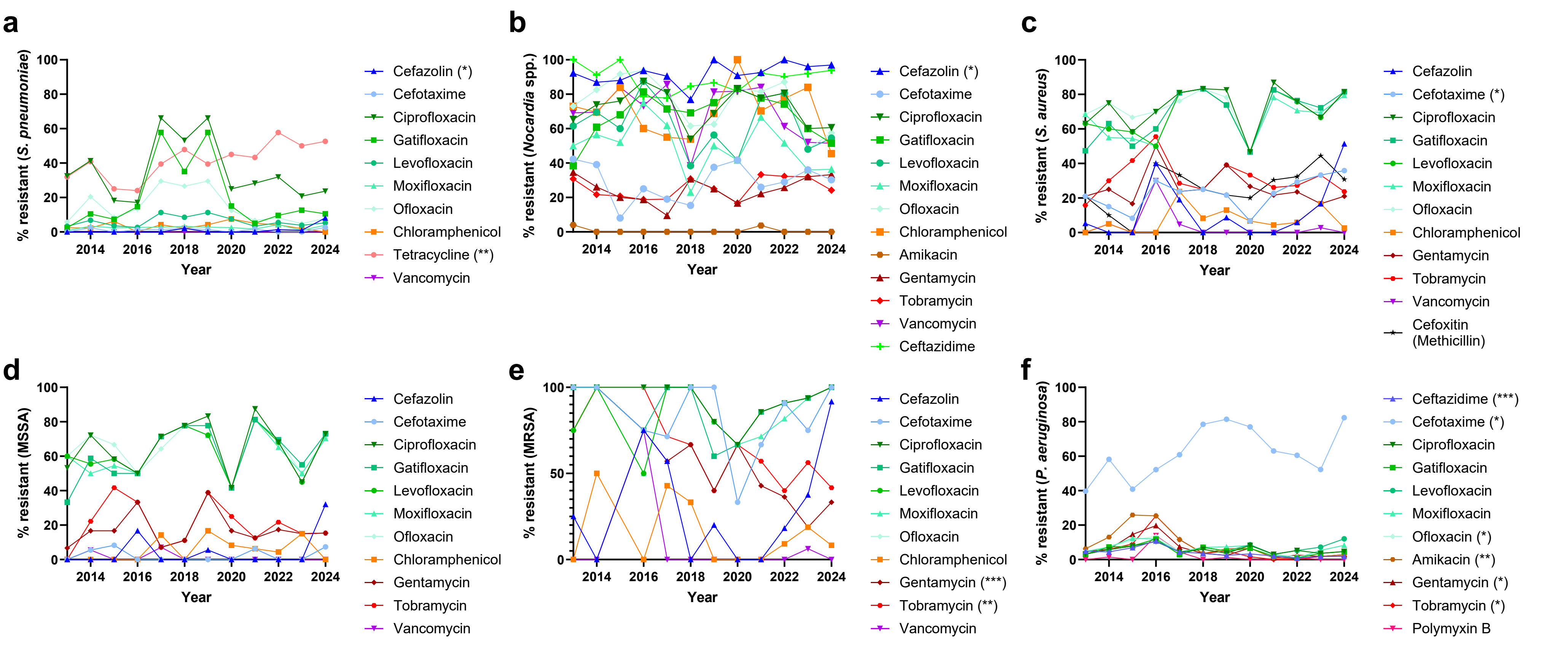

**Figure S1. Antimicrobial resistance rates of bacterial isolates cultured from corneal scrape samples 2013-2024.** Collected isolates underwent antimicrobial sensitivity testing. Data are presented as the % of all **(a)** *S. pneumoniae;* **(b)** *Nocardia* spp; **(c)** *S. aureus* (all); **(d)** methicillin sensitive *S. aureus* (MSSA); **(e)** methicillin resistant *S. aureus* (MRSA); and **(f)** *P. aeruginosa* isolates with resistance to antibiotics. Resistance determined by CLSI breakpoints (or those as outlined in the supplemental methodology section). Trends analysed by Spearman's rank correlation coefficient. *P ≤ 0.05; **P ≤ 0.01; *** P ≤ 0.001. The number of isolates evaluated per bacterial species are reported in supplementary tables S8-S13.

| **Supplementary Table S1. Antibiotic resistant breakpoints for each microorganism.** | | | | | |
|---|---|---|---|---|---|
| **Antibiotic class** | **Name of the antibiotic & disk content** | **Organism Name** | **Zone diameter breakpoint (mm)** | | |
| | | | Sensitive | Intermediate | Resistant |
| **Cephalosporins** | Cefazolin (30µg) | *Staphylococcus aureus* | ≥ 29 | - | ≤ 28 |
| | | *Streptococcus pneumoniae* | ≥ 29 | - | ≤ 28 |
| | | *Nocardia* spp. | ≥ 29 | - | ≤ 28 |
| | Cefotaxime (30µg) | *Staphylococcus aureus* | ≥ 29 | - | ≤ 28 |
| | | *Streptococcus pneumoniae* | ≥ 28 | 26-27 | ≤ 25 |
| | | *Nocardia* spp. | ≥ 29 | - | ≤ 28 |
| | | *Pseudomonas aeruginosa* | ≥ 26 | 23-25 | ≤ 22 |
| | Ceftazidime (30µg) | *Nocardia* spp. | ≥ 21 | 18-20 | ≤ 17 |
| | | *Pseudomonas aeruginosa* | ≥ 18 | 15-17 | ≤ 14 |
| | Cefoxitin (30µg) | *Staphylococcus aureus* | ≥ 22 | - | ≤ 21 |
| **Fluoroquinolones** | Levofloxacin (5µg) | *Staphylococcus aureus* | ≥ 19 | 16-18 | ≤ 15 |
| | | *Streptococcus pneumoniae* | ≥ 17 | 14-16 | ≤ 13 |
| | | *Nocardia* spp. | ≥ 19 | 16-18 | ≤ 15 |
| | | *Pseudomonas aeruginosa* | ≥ 22 | 15-21 | ≤ 14 |
| | Gatifloxacin (5µg) | *Staphylococcus aureus* | ≥ 23 | 20-22 | ≤ 19 |
| | | *Streptococcus pneumoniae* | ≥ 21 | 18-20 | ≤ 17 |
| | | *Nocardia* spp. | ≥ 23 | 20-22 | ≤ 19 |
| | | *Pseudomonas aeruginosa* | ≥ 18 | 15-17 | ≤ 14 |
| | Moxifloxacin (5µg) | *Staphylococcus aureus* | ≥ 24 | 21-23 | ≤ 20 |
| | | *Streptococcus pneumoniae* | ≥ 18 | 15-17 | ≤ 14 |
| | | *Nocardia* spp. | ≥ 24 | 21-23 | ≤ 20 |
| | | *Pseudomonas aeruginosa* | ≥ 18 | - | - |
| | Ciprofloxacin (5µg) | *Staphylococcus aureus* | ≥ 21 | 16-20 | ≤ 15 |
| | | *Streptococcus pneumoniae* | ≥ 21 | 16-20 | ≤ 15 |
| | | *Nocardia* spp. | ≥ 21 | 16-20 | ≤ 15 |
| | | *Pseudomonas aeruginosa* | ≥ 25 | 19-24 | ≤ 18 |
| | Ofloxacin (5µg) | *Staphylococcus aureus* | ≥ 18 | 15-17 | ≤ 14 |
| | | *Streptococcus pneumoniae* | ≥ 16 | 13-15 | ≤ 12 |
| | | *Nocardia* spp. | ≥ 18 | 15-17 | ≤ 14 |
| | | *Pseudomonas aeruginosa* | ≥16 | 13-15 | ≤ 12 |

**Supplementary Table S1. Antibiotic resistant breakpoints for each microorganism.**

| Antibiotic class | Name of the antibiotic & disk content | Organism Name | Zone diameter breakpoint (mm) | | |
|---|---|---|---|---|---|
| | | | Sensitive | Intermediate | Resistant |
| **Amphenicols** | Chloramphenicol (30 µg) | *Staphylococcus aureus* | ≥ 18 | 13-17 | ≤ 12 |
| | | *Streptococcus pneumoniae* | ≥ 21 | - | ≤ 20 |
| | | *Nocardia* spp. | ≥ 18 | 13-17 | ≤ 12 |
| **Aminoglycosides** | Gentamicin (10µg) | *Staphylococcus aureus* | ≥ 15 | 13-14 | ≤ 12 |
| | | *Nocardia* spp. | ≥ 15 | 13-14 | ≤ 12 |
| | | *Pseudomonas aeruginosa* | ≥ 15 | | ≤ 14 |
| | Tobramycin (10µg) | *Staphylococcus aureus* | ≥ 17 | 13-16 | ≤ 12 |
| | | *Nocardia* spp. | ≥ 17 | 13-16 | ≤ 12 |
| | | *Pseudomonas aeruginosa* | ≥ 19 | 13-18 | ≤ 12 |
| | Amikacin (30µg) | *Nocardia* spp. | ≥ 20 | 17-19 | ≤ 16 |
| | | *Pseudomonas aeruginosa* | ≥ 17 | 15-16 | ≤ 14 |
| **Glycopeptides** | Vancomycin (30µg) | *Staphylococcus aureus* | ≥ 17 | - | - |
| | | *Streptococcus pneumoniae* | ≥ 17 | - | - |
| | | *Nocardia* spp. | ≥ 17 | - | - |
| **Tetracyclines** | Tetracycline (30µg) | *Streptococcus pneumoniae* | ≥ 28 | 25-27 | ≤ 24 |
| **Polymyxins** | Polymyxin B (300units) | *Pseudomonas aeruginosa* | ≥ 12 | - | - |

**Supplementary Table S2. Summary of bacteria cultured from MK patients 2013-2024.**

| | | 2013 | 2014 | 2015 | 2016 | 2017 | 2018 | 2019 | 2020 | 2021 | 2022 | 2023 | 2024 | Total |
|---|---|---|---|---|---|---|---|---|---|---|---|---|---|---|
| **Gram-positive bacteria** | N | 287 | 258 | 222 | 164 | 215 | 191 | 208 | 125 | 192 | 250 | 266 | 249 | 2627 |
| | **(%)** | (71.9) | (69.7) | (62.4) | (61.9) | (66.4) | (57.4) | (57.6) | (57.6) | (56.8) | (60.7) | (58.7) | (59.0) | (61.81) |
| **Gram-negative bacteria** | **N** | 112 | 112 | 134 | 101 | 109 | 142 | 153 | 92 | 146 | 162 | 187 | 173 | 1623 |
| | **(%)** | (28.1) | (30.3) | (37.6) | (38.1) | (33.6) | (42.6) | (42.4) | (42.4) | (43.2) | (39.3) | (41.3) | (41.0) | (38.19) |
| Total | N | 399 | 370 | 356 | 265 | 324 | 333 | 361 | 217 | 338 | 412 | 453 | 422 | 4250 |

N= number of isolates. % = proportion of total isolates. Where significant (P<0.05) trends were found (by Spearman's rank correlation across entire data-set), category is shown in bold. Full statistical information shown in Supplementary Table S3.

**Supplementary Table S3. Summary of bacteria cultured from MK patients 2013-2024: Spearman's rank correlation coefficient.**

| | $r_s$ | 95% confidence interval | Strength of correlation | Significance (P) | |
|---|---|---|---|---|---|
| All bacteria (N) | 0.34 | -0.31 to 0.77 | weak | 0.2762 | ns |
| Gram-positive bacteria (N) | -0.08 | -0.64 to 0.53 | very weak | 0.8004 | ns |
| **Gram-negative bacteria (N)** | 0.68 | 0.15 to 0.90 | **strong** | **0.0187** | * |
| **Gram-positive bacteria (%)** | -0.66 | -0.90 to -0.11 | **strong** | **0.024** | * |
| **Gram-negative bacteria (%)** | 0.66 | 0.11 to 0.90 | **strong** | **0.024** | * |

N= number of isolates. % = proportion of total isolates. Where significant (P<0.05) trends were found (by Spearman's rank correlation across entire data-set), category is shown in bold.

### Supplementary Table S4. All gram-positive organisms cultured from MK patients 2013-2024.

| | | 2013 | 2014 | 2015 | 2016 | 2017 | 2018 | 2019 | 2020 | 2021 | 2022 | 2023 | 2024 | Total: |
|---|---|---|---|---|---|---|---|---|---|---|---|---|---|---|
| *Streptococcus pneumoniae* | N | 126 | 91 | 85 | 79 | 82 | 96 | 76 | 40 | 60 | 73 | 103 | 76 | 987 |
| | (%) | (43.9) | (35.3) | (38.3) | (48.2) | (38.1) | (50.3) | (36.5) | (32) | (31.3) | (29.2) | (38.7) | (30.5) | (37.57) |
| Other *Streptococcus* spp. | N | 5 | 6 | 2 | 1 | 4 | 1 | 11 | 4 | 2 | 4 | 2 | 7 | 49 |
| | (%) | (1.7) | (2.3) | (0.9) | (0.6) | (1.9) | (0.5) | (5.3) | (3.2) | (1) | (1.6) | (0.8) | (2.8) | (1.87) |
| *Corynebacterium* spp. | N | 39 | 49 | 39 | 26 | 34 | 29 | 28 | 17 | 18 | 32 | 42 | 40 | 393 |
| | (%) | (13.6) | (19) | (17.6) | (15.9) | (15.8) | (15.2) | (13.5) | (13.6) | (9.4) | (12.8) | (15.8) | (16.1) | (14.96) |
| ***Staphylococcus aureus*** | **N** | 22 | 22 | 14 | 13 | 25 | 16 | 26 | 19 | 23 | 34 | 36 | 40 | 290 |
| | **(%)** | (7.7) | (8.5) | (6.3) | (7.9) | (11.6) | (8.4) | (12.5) | (15.2) | (12) | (13.6) | (13.5) | (16.1) | (11.04) |
| *Nocardia* spp. | N | 27 | 23 | 27 | 17 | 21 | 13 | 16 | 12 | 28 | 32 | 25 | 35 | 276 |
| | (%) | (9.4) | (8.9) | (12.2) | (10.4) | (9.8) | (6.8) | (7.7) | (9.6) | (14.6) | (12.8) | (9.4) | (14.1) | (10.51) |
| Coagulase Negative Staphylococcus | N | 34 | 34 | 28 | 11 | 25 | 8 | 16 | 18 | 15 | 20 | 17 | 15 | 241 |
| | (%) | (11.8) | (13.2) | (12.6) | (6.7) | (11.6) | (4.2) | (7.7) | (14.4) | (7.8) | (8) | (6.4) | (6) | (9.17) |
| *Viridans streptococci* | N | 23 | 25 | 15 | 6 | 16 | 12 | 15 | 8 | 28 | 20 | 23 | 16 | 207 |
| | (%) | (8) | (9.7) | (6.8) | (3.7) | (7.4) | (6.3) | (7.2) | (6.4) | (14.6) | (8) | (8.6) | (6.4) | (7.88) |
| *Streptococcus pyogenes* | N | 5 | 3 | 3 | 4 | 3 | 5 | 2 | 0 | 5 | 2 | 2 | 4 | 38 |
| | (%) | (1.7) | (1.2) | (1.4) | (2.4) | (1.4) | (2.6) | (1) | (0) | (2.6) | (0.8) | (0.8) | (1.6) | (1.45) |
| Atypical *Mycobacterium* spp. | N | 2 | 0 | 7 | 1 | 1 | 6 | 7 | 2 | 5 | 4 | 4 | 4 | 43 |
| | (%) | (0.7) | (0) | (3.2) | (0.6) | (0.5) | (3.1) | (3.4) | (1.6) | (2.6) | (1.6) | (1.5) | (1.6) | (1.64) |
| ***Actinomyces* spp.** | **N** | 0 | 0 | 0 | 1 | 0 | 1 | 3 | 1 | 5 | 7 | 4 | 1 | 23 |
| | **(%)** | (0) | (0) | (0) | (0.6) | (0) | (0.5) | (1.4) | (0.8) | (2.6) | (2.8) | (1.5) | (0.4) | (0.88) |
| ***Propionibacterium acnes*** | **N** | 0 | 0 | 0 | 1 | 0 | 0 | 0 | 1 | 1 | 8 | 6 | 3 | 20 |
| | **(%)** | (0) | (0) | (0) | (0.6) | (0) | (0) | (0) | (0.8) | (0.5) | (3.2) | (2.3) | (1.2) | (0.76) |
| ***Bacillus* spp.** | **N** | 2 | 5 | 2 | 3 | 1 | 0 | 1 | 0 | 0 | 2 | 0 | 1 | 17 |
| | **(%)** | (0.7) | (1.9) | (0.9) | (1.8) | (0.5) | (0) | (0.5) | (0) | (0) | (0.8) | (0) | (0.4) | (0.65) |
| *Turicella otitidis* | N | 0 | 0 | 0 | 1 | 1 | 2 | 1 | 2 | 0 | 1 | 0 | 2 | 10 |
| | (%) | (0) | (0) | (0) | (0.6) | (0.5) | (1) | (0.5) | (1.6) | (0) | (0.4) | (0) | (0.8) | (0.38) |
| ***Kocuria kristinae*** | **N** | 0 | 0 | 0 | 0 | 0 | 0 | 1 | 1 | 0 | 2 | 1 | 2 | 7 |
| | **(%)** | (0) | (0) | (0) | (0) | (0) | (0) | (0.5) | (0.8) | (0) | (0.8) | (0.4) | (0.8) | (0.27) |
| | N | 1 | 0 | 0 | 0 | 0 | 1 | 0 | 0 | 1 | 2 | 1 | 0 | 6 |

| **Supplementary Table S4. All gram-positive organisms cultured from MK patients 2013-2024.** | | | | | | | | | | | | | | |
|---|---|---|---|---|---|---|---|---|---|---|---|---|---|---|
| | | **2013** | **2014** | **2015** | **2016** | **2017** | **2018** | **2019** | **2020** | **2021** | **2022** | **2023** | **2024** | **Total:** |
| *Eggerthia catenaformis* | (%) | (0.3) | (0) | (0) | (0) | (0) | (0.5) | (0) | (0) | (0.5) | (0.8) | (0.4) | (0) | (0.23) |
| ***Kocuria varians*** | **N** | 0 | 0 | 0 | 0 | 0 | 0 | 1 | 0 | 1 | 1 | 0 | 1 | 4 |
| | (%) | (0) | (0) | (0) | (0) | (0) | (0) | (0.5) | (0) | (0.5) | (0.4) | (0) | (0.4) | (0.15) |
| *Micrococcus* spp. | N | 1 | 0 | 0 | 0 | 2 | 0 | 1 | 0 | 0 | 0 | 0 | 0 | 4 |
| | (%) | (0.3) | (0) | (0) | (0) | (0.9) | (0) | (0.5) | (0) | (0) | (0) | (0) | (0) | (0.15) |
| *Arcanobacterium hemolyticum* | N | 0 | 0 | 0 | 0 | 0 | 1 | 0 | 0 | 0 | 1 | 0 | 0 | 2 |
| | (%) | (0) | (0) | (0) | (0) | (0) | (0.5) | (0) | (0) | (0) | (0.4) | (0) | (0) | (0.08) |
| *Kocuria rosea* | N | 0 | 0 | 0 | 0 | 0 | 0 | 0 | 0 | 0 | 0 | 0 | 2 | 2 |
| | (%) | (0) | (0) | (0) | (0) | (0) | (0) | (0) | (0) | (0) | (0) | (0) | (0.8) | (0.08) |
| *Lactobacillus hilgardii* | N | 0 | 0 | 0 | 0 | 0 | 0 | 1 | 0 | 0 | 1 | 0 | 0 | 2 |
| | (%) | (0) | (0) | (0) | (0) | (0) | (0) | (0.5) | (0) | (0) | (0.4) | (0) | (0) | (0.08) |
| *Rothia mucilaginosa* | N | 0 | 0 | 0 | 0 | 0 | 0 | 1 | 0 | 0 | 1 | 0 | 0 | 2 |
| | (%) | (0) | (0) | (0) | (0) | (0) | (0) | (0.5) | (0) | (0) | (0.4) | (0) | (0) | (0.08) |
| *Rothia dentocariosa* | N | 0 | 0 | 0 | 0 | 0 | 0 | 1 | 0 | 0 | 0 | 0 | 0 | 1 |
| | (%) | (0) | (0) | (0) | (0) | (0) | (0) | (0.5) | (0) | (0) | (0) | (0) | (0) | (0.04) |
| *Propionibacterium propionicum* | N | 0 | 0 | 0 | 0 | 0 | 0 | 0 | 0 | 0 | 1 | 0 | 0 | 1 |
| | (%) | (0) | (0) | (0) | (0) | (0) | (0) | (0) | (0) | (0) | (0.4) | (0) | (0) | (0.04) |
| *Atopobium vaginae* | N | 0 | 0 | 0 | 0 | 0 | 0 | 0 | 0 | 0 | 1 | 0 | 0 | 1 |
| | (%) | (0) | (0) | (0) | (0) | (0) | (0) | (0) | (0) | (0) | (0.4) | (0) | (0) | (0.04) |
| *Granulicatella adiacens* | N | 0 | 0 | 0 | 0 | 0 | 0 | 0 | 0 | 0 | 1 | 0 | 0 | 1 |
| | (%) | (0) | (0) | (0) | (0) | (0) | (0) | (0) | (0) | (0) | (0.4) | (0) | (0) | (0.04) |
| **Total** | | **287** | **258** | **222** | **164** | **215** | **191** | **208** | **125** | **192** | **250** | **266** | **249** | 2627 |
| (MSSA)^ | N | 15 | 18 | 12 | 6 | 14 | 9 | 18 | 12 | 16 | 23 | 20 | 27 | 190 |
| | (%) | (78.9) | (90.0) | (100.0) | (60.0) | (66.7) | (75.0) | (78.3) | (80.0) | (69.6) | (67.6) | (55.6) | (69.2) | (72.0) |
| **(MRSA)^** | **N** | 4 | 2 | 0 | 4 | 7 | 3 | 5 | 3 | 7 | 11 | 16 | 12 | 74 |
| | (%) | (21.1) | (10.0) | (0.0) | (40.0) | (33.3) | (25.0) | (21.7) | (20.0) | (30.4) | (32.4) | (44.4) | (30.8) | (28.0) |

N= number of isolates. % = proportion of gram-positive isolates. ^% proportion presented as total of *S. aureus* isolates. MSSA = methicillin sensitive *S. aureus*; MRSA = methicillin resistant *S. aureus*. Where significant (P<0.05) trends were found (by Spearman's rank correlation across entire data-set), category is shown in bold. Full statistical information shown in Supplementary Table S5.

**Supplementary Table S5. Gram-positive bacteria (N) Spearman's rank correlation coefficient.**

| | $r_s$ | 95% confidence interval | Strength of correlation | Trend (P) | |
|---|---|---|---|---|---|
| *Streptococcus pneumoniae* | -0.50 | -0.84 to 0.12 | moderate | 0.0996 | ns |
| Other *Streptococcus* spp. | 0.04 | -0.56 to 0.61 | very weak | 0.8973 | ns |
| *Corynebacterium* spp. | -0.12 | -0.66 to 0.50 | very weak | 0.7113 | ns |
| *Staphylococcus aureus* | 0.70 | 0.20 to 0.91 | **strong** | **0.0130** | * |
| *Nocardia* spp. | 0.29 | -0.35 to 0.75 | weak | 0.3503 | ns |
| *Coagulase Negative Staphylococcus* | -0.51 | -0.85 to 0.11 | moderate | 0.0912 | ns |
| *Viridans streptococci* | 0.07 | -0.54 to 0.63 | very weak | 0.8207 | ns |
| *Streptococcus pyogenes* | -0.31 | -0.76 to 0.34 | weak | 0.3272 | ns |
| *Atypical Mycobacterium* spp. | 0.32 | -0.33 to 0.76 | weak | 0.3105 | ns |
| *Actinomyces* spp. | 0.78 | 0.36 to 0.94 | **strong** | **0.0040** | ** |
| *Propionibacterium acnes* | 0.80 | 0.40 to 0.94 | **very strong** | **0.0032** | ** |
| *Bacillus* spp. | -0.61 | -0.88 to -0.031 | **strong** | **0.0403** | * |
| *Turicella otitidis* | 0.39 | -0.26 to 0.79 | weak | 0.2119 | ns |
| *Kocuria kristinae* | 0.79 | 0.38 to 0.94 | **strong** | **0.0045** | ** |
| *Eggerthia catenaformis* | 0.27 | -0.38 to 0.74 | weak | 0.3970 | ns |
| *Kocuria varians* | 0.61 | 0.043 to 0.88 | **strong** | **0.0485** | * |
| *Micrococcus* spp. | -0.35 | -0.78 to 0.30 | weak | 0.2697 | ns |
| *Arcanobacterium hemolyticum* | 0.19 | -0.44 to 0.70 | very weak | 0.6061 | ns |
| *Kocuria rosea* | 0.48 | -0.15 to 0.83 | moderate | 0.1667 | ns |
| *Lactobacillus hilgardii* | 0.26 | -0.39 to 0.73 | weak | 0.4848 | ns |
| *Rothia mucilaginosa* | 0.26 | -0.39 to 0.73 | weak | 0.4848 | ns |
| *Rothia dentocariosa* | 0.04 | -0.56 to 0.61 | very weak | >0.9999 | ns |
| *Propionibacterium propionicum* | 0.31 | -0.34 to 0.76 | very weak | 0.5000 | ns |
| *Atopobium vaginae* | 0.31 | -0.34 to 0.76 | very weak | 0.5000 | ns |
| *Granulicatella adiacens* | 0.31 | -0.34 to 0.76 | very weak | 0.5000 | ns |
| Methicillin sensitive *S. aureus* (MSSA) | 0.58 | -0.017 to 0.87 | moderate | 0.0537 | ns |
| Methicillin resistant *S. aureus* (MRSA) | 0.76 | 0.32 to 0.93 | **strong** | **0.0053** | ** |

**Supplementary Table S6. Gram-negative organisms cultured from MK patients 2013-2024.**

| | | 2013 | 2014 | 2015 | 2016 | 2017 | 2018 | 2019 | 2020 | 2021 | 2022 | 2023 | 2024 | Total: |
|---|---|---|---|---|---|---|---|---|---|---|---|---|---|---|
| ***Pseudomonas aeruginosa*** | **N** | 66 | 75 | 92 | 69 | 74 | 91 | 86 | 60 | 99 | 115 | 112 | 108 | 1047 |
| | (%) | (58.9) | (67) | (68.7) | (68.3) | (67.9) | (64.1) | (56.2) | (65.2) | (67.8) | (71) | (59.9) | (62.4) | (64.51) |
| Other *Pseudomonas* spp. | N | 3 | 4 | 6 | 3 | 3 | 3 | 11 | 3 | 7 | 3 | 4 | 3 | 53 |
| | (%) | (2.7) | (3.6) | (4.5) | (3) | (2.8) | (2.1) | (7.2) | (3.3) | (4.8) | (1.9) | (2.1) | (1.7) | (3.27) |
| ***Enterobacteriaceae* spp.** | N | 22 | 21 | 17 | 12 | 18 | 20 | 22 | 9 | 15 | 10 | 16 | 15 | 197 |
| | **(%)** | (19.6) | (18.8) | (12.7) | (11.9) | (16.5) | (14.1) | (14.4) | (9.8) | (10.3) | (6.2) | (8.6) | (8.7) | (12.14) |
| *Moraxella* spp. | N | 9 | 3 | 7 | 10 | 4 | 10 | 7 | 2 | 2 | 7 | 9 | 4 | 73 |
| | (%) | (8) | (2.7) | (5.2) | (9.9) | (3.7) | (7) | (4.6) | (2.2) | (1.4) | (4.3) | (4.8) | (2.3) | (4.50) |
| ***Aeromonas hydrophila*** | **N** | 0 | 0 | 0 | 0 | 0 | 0 | 0 | 7 | 6 | 9 | 7 | 13 | 42 |
| | (%) | (0) | (0) | (0) | (0) | (0) | (0) | (0) | (7.6) | (4.1) | (5.6) | (3.7) | (7.5) | (2.59) |
| **Other *Aeromonas* spp.** | **N** | 0 | 0 | 0 | 0 | 0 | 0 | 0 | 0 | 0 | 1 | 1 | 1 | 3 |
| | **(%)** | (0) | (0) | (0) | (0) | (0) | (0) | (0) | (0) | (0) | (0.6) | (0.5) | (0.6) | (0.18) |
| ***Acinetobacter baumannii* complex** | **N** | 0 | 1 | 0 | 0 | 3 | 2 | 6 | 1 | 6 | 1 | 9 | 3 | 32 |
| | **(%)** | (0) | (0.9) | (0) | (0) | (2.8) | (1.4) | (3.9) | (1.1) | (4.1) | (0.6) | (4.8) | (1.7) | (1.97) |
| Other *Acinetobacter* spp. | N | 0 | 1 | 4 | 3 | 0 | 5 | 3 | 0 | 3 | 1 | 2 | 2 | 24 |
| | (%) | (0) | (0.9) | (3) | (3) | (0) | (3.5) | (2) | (0) | (2.1) | (0.6) | (1.1) | (1.2) | (1.48) |
| ***Haemophilus* spp.** | **N** | 0 | 0 | 0 | 0 | 0 | 0 | 0 | 2 | 0 | 2 | 11 | 8 | 23 |
| | **(%)** | (0) | (0) | (0) | (0) | (0) | (0) | (0) | (2.2) | (0) | (1.2) | (5.9) | (4.6) | (1.42) |
| *Sphingomonas paucimobilis* | N | 0 | 0 | 0 | 1 | 1 | 2 | 0 | 1 | 3 | 7 | 2 | 1 | 18 |
| | (%) | (0) | (0) | (0) | (1) | (0.9) | (1.4) | (0) | (1.1) | (2.1) | (4.3) | (1.1) | (0.6) | (1.11) |
| *Burkholderia cepacia* | N | 3 | 0 | 1 | 0 | 0 | 2 | 3 | 2 | 1 | 1 | 2 | 2 | 17 |
| | (%) | (2.7) | (0) | (0.7) | (0) | (0) | (1.4) | (2) | (2.2) | (0.7) | (0.6) | (1.1) | (1.2) | (1.05) |
| *Neisseria* spp. | N | 0 | 0 | 0 | 0 | 3 | 3 | 6 | 0 | 0 | 1 | 0 | 2 | 15 |
| | (%) | (0) | (0) | (0) | (0) | (2.8) | (2.1) | (3.9) | (0) | (0) | (0.6) | (0) | (1.2) | (0.92) |
| **Unidentified gram-negative bacilli** | **N** | 3 | 6 | 2 | 0 | 0 | 1 | 0 | 0 | 0 | 0 | 0 | 0 | 12 |
| | **(%)** | (2.7) | (5.4) | (1.5) | (0) | (0) | (0.7) | (0) | (0) | (0) | (0) | (0) | (0) | (0.74) |
| | N | 1 | 0 | 0 | 0 | 0 | 2 | 2 | 1 | 0 | 1 | 3 | 1 | 11 |

| **Supplementary Table S6. Gram-negative organisms cultured from MK patients 2013-2024.** | | | | | | | | | | | | | | |
|---|---|---|---|---|---|---|---|---|---|---|---|---|---|---|
| | | **2013** | **2014** | **2015** | **2016** | **2017** | **2018** | **2019** | **2020** | **2021** | **2022** | **2023** | **2024** | **Total:** |
| *Stenotrophomonas maltophilia* | (%) | (0.9) | (0) | (0) | (0) | (0) | (1.4) | (1.3) | (1.1) | (0) | (0.6) | (1.6) | (0.6) | (0.68) |
| ***Achromobacter* spp.** | **N** | 0 | 0 | 0 | 1 | 0 | 1 | 0 | 1 | 1 | 2 | 1 | 4 | 11 |
| | **(%)** | (0) | (0) | (0) | (1) | (0) | (0.7) | (0) | (1.1) | (0.7) | (1.2) | (0.5) | (2.3) | (0.68) |
| *Capnocytophaga* spp. | N | 2 | 0 | 4 | 0 | 0 | 0 | 0 | 0 | 0 | 0 | 0 | 0 | 6 |
| | (%) | (1.8) | (0) | (3) | (0) | (0) | (0) | (0) | (0) | (0) | (0) | (0) | (0) | (0.37) |
| *Alcaligenes* spp. | N | 0 | 0 | 1 | 1 | 2 | 0 | 0 | 0 | 0 | 0 | 0 | 1 | 5 |
| | (%) | (0) | (0) | (0.7) | (1) | (1.8) | (0) | (0) | (0) | (0) | (0) | (0) | (0.6) | (0.31) |
| *Bordetella hinzii* | N | 0 | 0 | 0 | 0 | 1 | 0 | 0 | 0 | 0 | 0 | 2 | 0 | 3 |
| | (%) | (0) | (0) | (0) | (0) | (0.9) | (0) | (0) | (0) | (0) | (0) | (1.1) | (0) | (0.18) |
| *Comamonas testosteroni* | N | 0 | 0 | 0 | 0 | 0 | 0 | 0 | 0 | 0 | 0 | 3 | 0 | 3 |
| | (%) | (0) | (0) | (0) | (0) | (0) | (0) | (0) | (0) | (0) | (0) | (1.6) | (0) | (0.18) |
| *Pasteurella canis* | N | 0 | 0 | 0 | 1 | 0 | 0 | 0 | 0 | 0 | 0 | 0 | 2 | 3 |
| | (%) | (0) | (0) | (0) | (1) | (0) | (0) | (0) | (0) | (0) | (0) | (0) | (1.2) | (0.18) |
| *Ralstonia insidiosa* | N | 1 | 0 | 0 | 0 | 0 | 0 | 0 | 1 | 1 | 0 | 0 | 0 | 3 |
| | (%) | (0.9) | (0) | (0) | (0) | (0) | (0) | (0) | (1.1) | (0.7) | (0) | (0) | (0) | (0.18) |
| *Chryseobacterium ideologemes* | N | 0 | 0 | 0 | 0 | 0 | 0 | 0 | 0 | 0 | 1 | 1 | 0 | 2 |
| | (%) | (0) | (0) | (0) | (0) | (0) | (0) | (0) | (0) | (0) | (0.6) | (0.5) | (0) | (0.12) |
| *Cupriavidus pauculus* | N | 0 | 0 | 0 | 0 | 0 | 0 | 2 | 0 | 0 | 0 | 0 | 0 | 2 |
| | (%) | (0) | (0) | (0) | (0) | (0) | (0) | (1.3) | (0) | (0) | (0) | (0) | (0) | (0.12) |
| *Eikenella corrodens* | N | 1 | 0 | 0 | 0 | 0 | 0 | 0 | 1 | 0 | 0 | 0 | 0 | 2 |
| | (%) | (0.9) | (0) | (0) | (0) | (0) | (0) | (0) | (1.1) | (0) | (0) | (0) | (0) | (0.12) |
| *Ochrobactrum anthropi* | N | 0 | 0 | 0 | 0 | 0 | 0 | 1 | 1 | 0 | 0 | 0 | 0 | 2 |
| | (%) | (0) | (0) | (0) | (0) | (0) | (0) | (0.7) | (1.1) | (0) | (0) | (0) | (0) | (0.12) |
| *Ralstonia mannitolilytica* | N | 0 | 0 | 0 | 0 | 0 | 0 | 2 | 0 | 0 | 0 | 0 | 0 | 2 |
| | (%) | (0) | (0) | (0) | (0) | (0) | (0) | (1.3) | (0) | (0) | (0) | (0) | (0) | (0.12) |
| *Rhizobium radiobacter* | N | 0 | 0 | 0 | 0 | 0 | 0 | 0 | 0 | 1 | 0 | 0 | 1 | 2 |
| | (%) | (0) | (0) | (0) | (0) | (0) | (0) | (0) | (0) | (0.7) | (0) | (0) | (0.6) | (0.12) |
| *Brevundimonas diminuta* | N | 0 | 0 | 0 | 0 | 0 | 0 | 0 | 0 | 1 | 0 | 0 | 0 | 1 |
| | (%) | (0) | (0) | (0) | (0) | (0) | (0) | (0) | (0) | (0.7) | (0) | (0) | (0) | (0.06) |
| | N | 0 | 0 | 0 | 0 | 0 | 0 | 1 | 0 | 0 | 0 | 0 | 0 | 1 |

| **Supplementary Table S6. Gram-negative organisms cultured from MK patients 2013-2024.** | | | | | | | | | | | | | | |
|---|---|---|---|---|---|---|---|---|---|---|---|---|---|---|
| | | **2013** | **2014** | **2015** | **2016** | **2017** | **2018** | **2019** | **2020** | **2021** | **2022** | **2023** | **2024** | **Total:** |
| *Brevundimonas vesicularis* | (%) | (0) | (0) | (0) | (0) | (0) | (0) | (0.7) | (0) | (0) | (0) | (0) | (0) | (0.06) |
| *Delftia acidovorans* | N | 0 | 0 | 0 | 0 | 0 | 0 | 0 | 0 | 0 | 0 | 1 | 0 | 1 |
| | (%) | (0) | (0) | (0) | (0) | (0) | (0) | (0) | (0) | (0) | (0) | (0.5) | (0) | (0.06) |
| *Leclercia adecarboxylata* | N | 0 | 0 | 0 | 0 | 0 | 0 | 0 | 0 | 0 | 0 | 1 | 0 | 1 |
| | (%) | (0) | (0) | (0) | (0) | (0) | (0) | (0) | (0) | (0) | (0) | (0.5) | (0) | (0.06) |
| *Roseomonas gilardii* | N | 0 | 0 | 0 | 0 | 0 | 0 | 0 | 0 | 0 | 0 | 0 | 1 | 1 |
| | (%) | (0) | (0) | (0) | (0) | (0) | (0) | (0) | (0) | (0) | (0) | (0) | (0.6) | (0.06) |
| *Shewanella algae* | N | 0 | 0 | 0 | 0 | 0 | 0 | 0 | 0 | 0 | 0 | 0 | 1 | 1 |
| | (%) | (0) | (0) | (0) | (0) | (0) | (0) | (0) | (0) | (0) | (0) | (0) | (0.6) | (0.06) |
| *Shewanella amazonensis* | N | 1 | 0 | 0 | 0 | 0 | 0 | 0 | 0 | 0 | 0 | 0 | 0 | 1 |
| | (%) | (0.9) | (0) | (0) | (0) | (0) | (0) | (0) | (0) | (0) | (0) | (0) | (0) | (0.06) |
| *Shewanella putrefaciens* | N | 0 | 1 | 0 | 0 | 0 | 0 | 0 | 0 | 0 | 0 | 0 | 0 | 1 |
| | (%) | (0) | (0.9) | (0) | (0) | (0) | (0) | (0) | (0) | (0) | (0) | (0) | (0) | (0.06) |
| *Sphingomonas thalpophilum* | N | 0 | 0 | 0 | 0 | 0 | 0 | 1 | 0 | 0 | 0 | 0 | 0 | 1 |
| | (%) | (0) | (0) | (0) | (0) | (0) | (0) | (0.7) | (0) | (0) | (0) | (0) | (0) | (0.06) |
| Total | | 112 | 112 | 134 | 101 | 109 | 142 | 153 | 92 | 146 | 162 | 187 | 173 | 1047 |

N= number of isolates. % = proportion of gram-negative isolates. Where significant (P<0.05) trends were found (by Spearman's rank correlation across entire data-set), category is shown in bold. Full statistical information shown in Supplementary Table S7.

| **Supplementary Table S7. Gram-negative bacteria (n) Spearman's rank correlation coefficient.** | | | | | |
|---|---|---|---|---|---|
| | $r_s$ | 95% confidence interval | Strength of correlation | P | |
| *Pseudomonas aeruginosa* | 0.66 | 0.13 to 0.90 | **strong** | **0.0219** | * |
| Other *Pseudomonas* spp. | 0.00 | -0.58 to 0.59 | very weak | 0.9964 | ns |
| *Enterobacteriaceae* spp. | -0.56 | -0.86 to 0.043 | moderate | 0.0628 | ns |
| *Moraxella* spp. | -0.21 | -0.71 to 0.43 | weak | 0.5038 | ns |
| *Aeromonas hydrophila* | 0.87 | 0.58 to 0.96 | very strong | **0.0003** | *** |
| *Other Aeromonas* spp. | 0.75 | 0.30 to 0.93 | **strong** | **0.0091** | ** |
| *Acinetobacter baumannii complex* | 0.68 | 0.15 to 0.91 | **strong** | **0.0181** | * |
| *Other Acinetobacter* spp. | 0.07 | -0.54 to 0.63 | very weak | 0.8355 | ns |
| *Haemophilus* spp. | 0.79 | 0.38 to 0.94 | **strong** | **0.0037** | ** |
| *Sphingomonas paucimobilis* | 0.69 | 0.17 to 0.91 | **strong** | **0.0167** | * |
| *Burkholderia cepacia* | 0.23 | -0.41 to 0.72 | weak | 0.4727 | ns |
| *Neisseria* spp. | 0.25 | -0.39 to 0.73 | weak | 0.4225 | ns |
| *Unidentified gram-negative bacilli* | -0.76 | -0.93 to -0.31 | **strong** | **0.0064** | ** |
| *Stenotrophomonas maltophilia* | 0.47 | -0.17 to 0.83 | moderate | 0.1305 | ns |
| *Achromobacter* spp. | 0.79 | 0.38 to 0.94 | **strong** | **0.0035** | ** |
| *Capnocytophaga* spp. | -0.57 | -0.87 to 0.025 | moderate | 0.0606 | ns |
| *Alcaligenes* spp. | -0.12 | -0.66 to 0.50 | very weak | 0.7192 | ns |
| *Bordetella hinzii* | 0.23 | -0.42 to 0.72 | weak | 0.5 | ns |
| *Comamonas testosteroni* | 0.39 | -0.25 to 0.80 | weak | 0.3333 | ns |
| *Pasteurella canis* | 0.24 | -0.41 to 0.72 | weak | 0.4697 | ns |
| *Ralstonia insidiosa* | -0.08 | -0.64 to 0.53 | very weak | 0.8636 | ns |
| *Chryseobacterium ideologemes* | 0.52 | -0.098 to 0.85 | moderate | 0.1212 | ns |

| **Supplementary Table S7. Gram-negative bacteria (n) Spearman's rank correlation coefficient.** | | | | | |
|---|---|---|---|---|---|
| | **$r_s$** | **95% confidence interval** | **Strength of correlation** | **P** | |
| *Cupriavidus pauculus* | 0.04 | -0.56 to 0.61 | very weak | >0.9999 | ns |
| *Eikenella corrodens* | -0.26 | -0.73 to 0.39 | weak | 0.4848 | ns |
| *Ochrobactrum anthropi* | 0.13 | -0.49 to 0.67 | very weak | 0.7576 | ns |
| *Ralstonia mannitolilytica* | 0.04 | -0.56 to 0.61 | very weak | >0.9999 | ns |
| *Rhizobium radiobacter* | 0.52 | -0.098 to 0.85 | moderate | 0.1212 | ns |
| *Brevundimonas diminuta* | 0.22 | -0.42 to 0.71 | weak | 0.6667 | ns |
| *Brevundimonas vesicularis* | 0.04 | -0.56 to 0.61 | very weak | >0.9999 | ns |
| *Delftia acidovorans* | 0.39 | -0.25 to 0.80 | weak | 0.3333 | ns |
| *Leclercia adecarboxylata* | 0.39 | -0.25 to 0.80 | weak | 0.3333 | ns |
| *Roseomonas gilardii* | 0.48 | -0.15 to 0.83 | moderate | 0.1667 | ns |
| *Shewanella algae* | 0.48 | -0.15 to 0.83 | moderate | 0.1667 | ns |
| *Shewanella amazonensis* | -0.48 | -0.83 to 0.15 | moderate | 0.1667 | ns |
| *Shewanella putrefaciens* | -0.39 | -0.80 to 0.25 | weak | 0.3333 | ns |
| *Sphingomonas thalpophilum* | 0.04 | -0.56 to 0.61 | weak | >0.9999 | ns |

**Supplementary Table S8: Number of isolates and resistance profiles of *S. pneumoniae* (SP) per year.**

| | | | 2013 | 2014 | 2015 | 2016 | 2017 | 2018 | 2019 | 2020 | 2021 | 2022 | 2023 | 2024 | All years |
|---|---|---|---|---|---|---|---|---|---|---|---|---|---|---|---|
| | | *N=* | *123* | *88* | *82* | *76* | *71* | *94* | *71* | *40* | *60* | *73* | *103* | *76* | *957* |
| | **Antibiotic class** | **Antibiotic** | **% of isolates with resistance** | | | | | | | | | | | | |
| **SP** | Cephalosporins | **Cefazolin** | 0.0 | 0.0 | 0.0 | 0.0 | 0.0 | 2.1 | 0.0 | 0.0 | 0.0 | 1.4 | 1.0 | 8.1 | 1.1 |
| | | Cefotaxime | 0.0 | 2.4 | 0.0 | 1.3 | 1.4 | 2.1 | 1.4 | 0.0 | 1.7 | 0.0 | 0.0 | 2.6 | 1.1 |
| | Fluoroquinolones | Levofloxacin | 3.3 | 6.8 | 3.7 | 2.6 | 11.3 | 8.5 | 11.3 | 7.5 | 3.3 | 5.5 | 3.9 | 5.3 | 5.9 |
| | | Gatifloxacin | 2.4 | 10.5 | 7.4 | 14.7 | 57.7 | 35.1 | 57.7 | 15.0 | 5.0 | 9.6 | 12.6 | 10.5 | 19.0 |
| | | Moxifloxacin | 0.8 | 3.4 | 2.4 | 2.6 | 2.8 | 3.2 | 2.8 | 2.5 | 1.7 | 4.3 | 1.0 | 3.9 | 2.5 |
| | | Ciprofloxacin | 32.5 | 41.4 | 18.3 | 17.1 | 66.2 | 53.2 | 66.2 | 25.0 | 28.3 | 31.9 | 20.8 | 23.7 | 35.4 |
| | | Ofloxacin | 5.7 | 20.5 | 8.5 | 13.3 | 29.6 | 26.6 | 29.6 | 12.5 | 6.7 | 8.2 | 4.9 | 7.9 | 14.1 |
| | Amphenicols | Chloramphenicol | 2.5 | 2.3 | 6.2 | 0.0 | 4.2 | 2.1 | 4.2 | 7.5 | 5.1 | 4.1 | 2.0 | 0.0 | 3.0 |
| | Tetracyclines | **Tetracycline** | 32.0 | 40.9 | 25.0 | 24.0 | 39.4 | 47.9 | 39.4 | 45.0 | 43.3 | 57.7 | 50.0 | 52.6 | 41.1 |
| | Glycopeptides | Vancomycin | 0.0 | 0.0 | 0.0 | 0.0 | 0.0 | 0.0 | 0.0 | 0.0 | 0.0 | 0.0 | 0.0 | 0.0 | 0.0 |

P values calculated from Spearman's rank correlation across entire data-set. Antibiotics with significant (P<0.05) trends over the time course are depicted in bold. Full statistical analysis provided in Table 1.

**Supplementary Table S9: Number of isolates and resistance profiles of *Nocardia* spp. (No) per year.**

| | | | 2013 | 2014 | 2015 | 2016 | 2017 | 2018 | 2019 | 2020 | 2021 | 2022 | 2023 | 2024 | All years |
|---|---|---|---|---|---|---|---|---|---|---|---|---|---|---|---|
| | | *N=* | *26* | *23* | *25* | *16* | *21* | *13* | *16* | *12* | *27* | *31* | *25* | *33* | *268* |
| | **Antibiotic class** | **Antibiotic** | **% of isolates with resistance** | | | | | | | | | | | | |
| **No** | Cephalosporins | **Cefazolin** | 92.3 | 87.0 | 88.0 | 93.8 | 90.5 | 76.9 | 100.0 | 90.9 | 92.6 | 100.0 | 96.0 | 97.0 | 92.9 |
| | | Cefotaxime | 42.3 | 39.1 | 8.0 | 25.0 | 19.0 | 15.4 | 37.5 | 41.7 | 25.9 | 29.0 | 36.0 | 30.3 | 29.1 |
| | | Ceftazidime | 100.0 | 91.3 | 100.0 | 78.6 | 77.8 | 84.6 | 86.7 | 81.8 | 92.3 | 90.3 | 92.0 | 93.9 | 90.7 |
| | Fluoroquinolones | Levofloxacin | 61.5 | 69.6 | 60.0 | 87.5 | 71.4 | 38.5 | 56.3 | 41.7 | 77.8 | 80.6 | 48.0 | 54.5 | 63.8 |
| | | Gatifloxacin | 38.5 | 60.9 | 68.0 | 81.3 | 71.4 | 69.2 | 75.0 | 83.3 | 77.8 | 74.2 | 60.0 | 51.5 | 65.7 |
| | | Moxifloxacin | 50.0 | 56.5 | 52.0 | 75.0 | 61.9 | 23.1 | 50.0 | 41.7 | 66.7 | 51.6 | 36.0 | 36.4 | 50.4 |
| | | Ciprofloxacin | 65.4 | 73.9 | 76.0 | 87.5 | 81.0 | 53.8 | 68.8 | 83.3 | 77.8 | 80.6 | 60.0 | 60.6 | 72.0 |
| | | Ofloxacin | 73.1 | 82.6 | 91.7 | 93.8 | 81.0 | 61.5 | 62.5 | 83.3 | 81.5 | 87.1 | 48.0 | 57.6 | 74.9 |
| | Amphenicols | Chloramphenicol | 73.1 | 69.6 | 84.0 | 60.0 | 55.0 | 53.8 | 68.8 | 100.0 | 70.4 | 77.4 | 84.0 | 45.5 | 69.5 |
| | Aminoglycosides | Amikacin | 4.0 | 0.0 | 0.0 | 0.0 | 0.0 | 0.0 | 0.0 | 0.0 | 3.7 | 0.0 | 0.0 | 0.0 | 0.8 |
| | | Gentamicin | 34.6 | 26.1 | 20.0 | 18.8 | 9.5 | 30.8 | 25.0 | 16.7 | 22.2 | 25.8 | 32.0 | 33.3 | 25.4 |
| | | Tobramycin | 30.8 | 21.7 | 20.8 | 18.8 | 19.0 | 30.8 | 25.0 | 16.7 | 33.3 | 32.3 | 32.0 | 24.2 | 26.2 |
| | Glycopeptides | Vancomycin | 69.2 | 69.6 | 84.0 | 73.3 | 85.7 | 38.5 | 81.3 | 81.8 | 84.0 | 61.3 | 52.0 | 51.5 | 68.6 |

P values calculated from Spearman's rank correlation across entire data-set. Antibiotics with significant (P<0.05) trends over the time course are depicted in bold. Full statistical analysis provided in Table 1.

**Supplementary Table S10: Number of isolates and resistance profiles of *S. aureus* (SA) per year.**

| | | | 2013 | 2014 | 2015 | 2016 | 2017 | 2018 | 2019 | 2020 | 2021 | 2022 | 2023 | 2024 | All years |
|---|---|---|---|---|---|---|---|---|---|---|---|---|---|---|---|
| | | *N=* | *19* | *20* | *12* | *10* | *21* | *12* | *23* | *15* | *23* | *34* | *36* | *39* | *264* |
| | **Antibiotic class** | **Antibiotic** | **% of isolates with resistance** | | | | | | | | | | | | |
| **SA (all)** | Cephalosporins | Cefazolin | 5.3 | 0.0 | 0.0 | 40.0 | 19.0 | 0.0 | 8.7 | 0.0 | 0.0 | 5.9 | 16.7 | 51.4 | 14.5 |
| | | **Cefotaxime** | 21.1 | 15.0 | 8.3 | 30.0 | 23.8 | 25.0 | 21.7 | 6.7 | 22.7 | 29.4 | 33.3 | 35.9 | 25.1 |
| | | Cefoxitin | 21.1 | 10.0 | 0.0 | 40.0 | 33.3 | 25.0 | 21.7 | 20.0 | 30.4 | 32.4 | 44.4 | 30.8 | 28.0 |
| | Fluoroquinolones | Levofloxacin | 63.2 | 60.0 | 58.3 | 50.0 | 81.0 | 83.3 | 73.9 | 46.7 | 82.6 | 75.8 | 66.7 | 81.6 | 71.0 |
| | | Gatifloxacin | 47.4 | 63.2 | 50.0 | 60.0 | 81.0 | 83.3 | 73.9 | 46.7 | 82.6 | 76.5 | 72.2 | 81.6 | 71.0 |
| | | Moxifloxacin | 68.4 | 55.0 | 54.5 | 50.0 | 81.0 | 83.3 | 73.9 | 46.7 | 78.3 | 70.6 | 69.4 | 79.5 | 70.0 |
| | | Ciprofloxacin | 63.2 | 75.0 | 58.3 | 70.0 | 81.0 | 83.3 | 82.6 | 46.7 | 87.0 | 75.8 | 66.7 | 81.6 | 74.0 |
| | | Ofloxacin | 68.4 | 75.0 | 66.7 | 70.0 | 76.2 | 83.3 | 78.3 | 46.7 | 82.6 | 76.5 | 69.4 | 79.5 | 73.9 |
| | Amphenicols | Chloramphenicol | 0.0 | 5.0 | 0.0 | 0.0 | 23.8 | 8.3 | 13.0 | 6.7 | 4.3 | 5.9 | 16.7 | 2.6 | 8.0 |
| | Aminoglycosides | Gentamicin | 21.1 | 25.0 | 16.7 | 50.0 | 23.8 | 25.0 | 39.1 | 26.7 | 21.7 | 23.5 | 16.7 | 21.1 | 24.3 |
| | | Tobramycin | 15.8 | 30.0 | 41.7 | 55.6 | 28.6 | 25.0 | 39.1 | 33.3 | 26.1 | 27.3 | 33.3 | 23.7 | 29.9 |
| | Glycopeptides | Vancomycin | 0.0 | 5.0 | 0.0 | 30.0 | 4.8 | 0.0 | 0.0 | 0.0 | 0.0 | 0.0 | 2.8 | 0.0 | 2.3 |

P values calculated from Spearman's rank correlation across entire data-set. Antibiotics with significant (P<0.05) trends over the time course are depicted in bold. Full statistical analysis provided in Table 1.

**Supplementary Table S11: Number of isolates and resistance profiles of *S. aureus* (SA): MSSA per year.**

| | | | 2013 | 2014 | 2015 | 2016 | 2017 | 2018 | 2019 | 2020 | 2021 | 2022 | 2023 | 2024 | All years |
|---|---|---|---|---|---|---|---|---|---|---|---|---|---|---|---|
| | | *N=* | *15* | *18* | *12* | *6* | *14* | *9* | *18* | *12* | *16* | *23* | *20* | *27* | *190* |
| | **Antibiotic class** | **Antibiotic** | **% of isolates with resistance** | | | | | | | | | | | | |
| **SA (MSSA)** | Cephalosporins | Cefazolin | 0.0 | 0.0 | 0.0 | 16.7 | 0.0 | 0.0 | 5.6 | 0.0 | 0.0 | 0.0 | 0.0 | 32.0 | 5.3 |
| | | Cefotaxime | 0.0 | 5.6 | 8.3 | 0.0 | 0.0 | 0.0 | 0.0 | 0.0 | 6.3 | 0.0 | 0.0 | 7.4 | 2.6 |
| | | Cefoxitin | 0.0 | 0.0 | 0.0 | 0.0 | 0.0 | 0.0 | 0.0 | 0.0 | 0.0 | 0.0 | 0.0 | 0.0 | 0.0 |
| | Fluoroquinolones | Levofloxacin | 60.0 | 55.6 | 58.3 | 50.0 | 71.4 | 77.8 | 72.2 | 41.7 | 81.3 | 68.2 | 45.0 | 73.1 | 63.8 |
| | | Gatifloxacin | 33.3 | 58.8 | 50.0 | 50.0 | 71.4 | 77.8 | 77.8 | 41.7 | 81.3 | 69.6 | 55.0 | 73.1 | 63.3 |
| | | Moxifloxacin | 60.0 | 50.0 | 54.5 | 50.0 | 71.4 | 77.8 | 72.2 | 41.7 | 81.3 | 65.2 | 50.0 | 70.4 | 63.0 |
| | | Ciprofloxacin | 53.3 | 72.2 | 58.3 | 50.0 | 71.4 | 77.8 | 83.3 | 41.7 | 87.5 | 68.2 | 45.0 | 73.1 | 66.5 |
| | | Ofloxacin | 60.0 | 72.2 | 66.7 | 50.0 | 64.3 | 77.8 | 77.8 | 41.7 | 81.3 | 69.6 | 50.0 | 70.4 | 66.3 |
| | Amphenicols | Chloramphenicol | 0.0 | 0.0 | 0.0 | 0.0 | 14.3 | 0.0 | 16.7 | 8.3 | 6.3 | 4.3 | 15.0 | 0.0 | 5.8 |
| | Aminoglycosides | Gentamicin | 6.7 | 16.7 | 16.7 | 33.3 | 7.1 | 11.1 | 38.9 | 16.7 | 12.5 | 17.4 | 15.0 | 15.4 | 16.9 |
| | | Tobramycin | 0.0 | 22.2 | 41.7 | 33.3 | 7.1 | 11.1 | 38.9 | 25.0 | 12.5 | 21.7 | 15.0 | 15.4 | 19.6 |
| | Glycopeptides | Vancomycin | 0.0 | 5.6 | 0.0 | 0.0 | 7.1 | 0.0 | 0.0 | 0.0 | 0.0 | 0.0 | 0.0 | 0.0 | 1.1 |

P values calculated from Spearman's rank correlation across entire data-set. Antibiotics with significant (P<0.05) trends over the time course are depicted in bold. Full statistical analysis provided in Table 1.

**Supplementary Table S12: Number of isolates and resistance profiles of *S. aureus* (SA): MRSA per year.**

| | | | 2013 | 2014 | 2015 | 2016 | 2017 | 2018 | 2019 | 2020 | 2021 | 2022 | 2023 | 2024 | All years |
|---|---|---|---|---|---|---|---|---|---|---|---|---|---|---|---|
| | | *N=* | *4* | *2* | *0* | *4* | *7* | *3* | *5* | *3* | *7* | *11* | *16* | *12* | *74* |
| | **Antibiotic class** | **Antibiotic** | **% of isolates with resistance** | | | | | | | | | | | | |
| **SA (MRSA)** | Cephalosporins | Cefazolin | 25.0 | 0.0 | N/A | 75.0 | 57.1 | 0.0 | 20.0 | 0.0 | 0.0 | 18.2 | 37.5 | 91.7 | 37.8 |
| | | Cefotaxime | 100.0 | 100.0 | N/A | 75.0 | 71.4 | 100.0 | 100.0 | 33.3 | 66.7 | 90.9 | 75.0 | 100.0 | 83.6 |
| | | Cefoxitin | 100.0 | 100.0 | N/A | 100.0 | 100.0 | 100.0 | 100.0 | 100.0 | 100.0 | 100.0 | 100.0 | 100.0 | 100.0 |
| | Fluoroquinolones | Levofloxacin | 75.0 | 100.0 | N/A | 50.0 | 100.0 | 100.0 | 80.0 | 66.7 | 85.7 | 90.9 | 93.8 | 100.0 | 89.2 |
| | | Gatifloxacin | 100.0 | 100.0 | N/A | 75.0 | 100.0 | 100.0 | 60.0 | 66.7 | 85.7 | 90.9 | 93.8 | 100.0 | 90.5 |
| | | Moxifloxacin | 100.0 | 100.0 | N/A | 50.0 | 100.0 | 100.0 | 80.0 | 66.7 | 71.4 | 81.8 | 93.8 | 100.0 | 87.8 |
| | | Ciprofloxacin | 100.0 | 100.0 | N/A | 100.0 | 100.0 | 100.0 | 80.0 | 66.7 | 85.7 | 90.9 | 93.8 | 100.0 | 93.2 |
| | | Ofloxacin | 100.0 | 100.0 | N/A | 100.0 | 100.0 | 100.0 | 80.0 | 66.7 | 85.7 | 90.9 | 93.8 | 100.0 | 93.2 |
| | Amphenicols | Chloramphenicol | 0.0 | 50.0 | N/A | 0.0 | 42.9 | 33.3 | 0.0 | 0.0 | 0.0 | 9.1 | 18.8 | 8.3 | 13.5 |
| | Aminoglycosides | **Gentamicin** | 75.0 | 100.0 | N/A | 75.0 | 57.1 | 66.7 | 40.0 | 66.7 | 42.9 | 36.4 | 18.8 | 33.3 | 43.2 |
| | | **Tobramycin** | 75.0 | 100.0 | N/A | 100.0 | 71.4 | 66.7 | 40.0 | 66.7 | 57.1 | 40.0 | 56.3 | 41.7 | 56.9 |
| | Glycopeptides | Vancomycin | 0.0 | 0.0 | N/A | 75.0 | 0.0 | 0.0 | 0.0 | 0.0 | 0.0 | 0.0 | 6.3 | 0.0 | 5.4 |

P values calculated from Spearman's rank correlation across entire data-set. Antibiotics with significant (P<0.05) trends over the time course are depicted in bold. Full statistical analysis provided in Table 1.

**Supplementary Table S13: Number of isolates and resistance profiles of *P. aeruginosa* (PA) per year.**

| | | | 2013 | 2014 | 2015 | 2016 | 2017 | 2018 | 2019 | 2020 | 2021 | 2022 | 2023 | 2024 | All years |
|---|---|---|---|---|---|---|---|---|---|---|---|---|---|---|---|
| | | *N=* | *64* | *69* | *89* | *67* | *69* | *84* | *81* | *61* | *100* | *114* | *111* | *108* | *1017* |
| | **Antibiotic class** | **Antibiotic** | **% of isolates with resistance** | | | | | | | | | | | | |
| **PA** | Cephalosporins | **Cefotaxime** | 39.7 | 58.2 | 40.9 | 52.2 | 60.9 | 78.6 | 81.5 | 77 | 63 | 60.5 | 52.3 | 82.4 | 62.7 |
| | | **Ceftazidime** | 4.8 | 4.5 | 6.9 | 10.8 | 4.4 | 3.6 | 2.5 | 3.3 | 2.0 | 0.9 | 1.8 | 1.9 | 3.6 |
| | Fluoroquinolones | Levofloxacin | 3.2 | 5.8 | 7.9 | 11.9 | 2.9 | 7.1 | 4.9 | 8.6 | 3.0 | 5.3 | 7.2 | 12.0 | 6.7 |
| | | Gatifloxacin | 3.1 | 7.2 | 7.9 | 11.9 | 2.9 | 7.1 | 4.9 | 6.6 | 2.0 | 0.9 | 3.6 | 4.6 | 4.9 |
| | | Moxifloxacin | 4.7 | 7.2 | 12.4 | 11.9 | 4.3 | 7.1 | 7.4 | 8.2 | 3.0 | 0.9 | 5.4 | 8.3 | 6.5 |
| | | Ciprofloxacin | 3.2 | 5.8 | 7.9 | 10.4 | 4.3 | 6.0 | 3.7 | 8.2 | 3.0 | 5.3 | 3.6 | 4.7 | 5.3 |
| | | **Ofloxacin** | 4.7 | 7.2 | 11.4 | 13.4 | 4.3 | 7.1 | 4.9 | 6.6 | 3.0 | 1.8 | 3.6 | 4.6 | 5.7 |
| | Aminoglycosides | **Amikacin** | 6.3 | 13.0 | 25.8 | 25.4 | 11.6 | 3.6 | 6.2 | 6.6 | 1.0 | 0.9 | 1.8 | 2.8 | 7.9 |
| | | **Gentamicin** | 4.8 | 5.9 | 14.6 | 19.7 | 7.4 | 3.6 | 2.5 | 6.7 | 1.0 | 2.6 | 1.8 | 2.8 | 5.5 |
| | | **Tobramycin** | 3.2 | 5.9 | 9.1 | 25.4 | 11.6 | 3.6 | 4.9 | 1.6 | 0.0 | 0.0 | 1.8 | 1.9 | 5.1 |
| | Polymyxins | Polymyxin B | 0.0 | 1.5 | 0.0 | 13.8 | 4.4 | 0.0 | 1.3 | 0.0 | 0.0 | 0.9 | 0.0 | 0.0 | 1.5 |

P values calculated from Spearman's rank correlation across entire data-set. Antibiotics with significant ($P<0.05$) trends over the time course are depicted in bold. Full statistical analysis provided in Table 1.
