## Supplemental materials and methods for "Trends in aetiology and antibiotic resistance of bacterial keratitis isolates from South India between 2013-2024"

1. **Microbiological identification of pathogens**
2. **Antibacterial susceptibility testing**
3. **Spearman's rank correlation coefficient ($R_s$) interpretation**

1. **Microbiological identification of pathogens**

Following thorough slit-lamp biomicroscopic examination, corneal scraping specimens were collected by an ophthalmologist by standard clinical procedure for both direct microscopic observation and culture. The collected samples were inoculated onto 5% sheep blood agar and potato dextrose agar. Specimens inoculated onto blood agar were incubated at 37°C in 5% $CO_2$ for 24 h and those onto potato dextrose agar were incubated at 25–27°C for up to two weeks. Following growth, pure cultures of the organisms were used for species level identification through standard microscopic and biochemical tests.

2. **Antibacterial susceptibility testing**

The antibacterial agents used were: amikacin (30 µg), cefazolin (30 µg), cefotaxime (30 µg), ceftazidime (30 µg), chloramphenicol (30 µg), ciprofloxacin (5 µg), gatifloxacin (5 µg), gentamicin (10 µg), levofloxacin (5 µg), moxifloxacin (5 µg), ofloxacin (5 µg), polymyxin B (300 U), tetracycline (30µg), tobramycin (10 µg), and vancomycin (30 µg) as appropriate per bacterial species. Cefoxitin (30 µg) was used as a surrogate marker for methicillin resistance, acting as an inducer of the mecA gene (1).

The assay results were interpreted in accordance with the Clinical and Laboratory Standards Institute guidelines (CLSI) (2) or from the European Committee on Antimicrobial Susceptibility testing (EUCAST) guidelines (3) if CSLI guidelines were not available.

Where neither were available, the following breakpoints were adopted:
For *P. aeruginosa,* breakpoints for *Enterobacteriaceae* spp. against gatifloxacin were used. Currently, CLSI and EUCAST do not provide the breakpoints for polymyxin-B by disc diffusion, only MIC breakpoints are available. However, since polymyxin-B still continues demonstrate good clinical efficacy, testing by disk diffusion is still performed and results interpreted using the previously reported breakpoints.

For *Nocardia* spp. there are no CLSI resistance breakpoints for the disc diffusion assay, therefore, we used the same breakpoints that we used for *Staphylococcus aureus*.

CLSI removed the cefazolin breakpoints for *Staphylococcus* spp. in 2013, however, we continued to follow the older breakpoints (susceptible ≥18 mm, intermediate 15–17 mm, resistant ≤14 mm) until 2021. In 2022, we updated our lab manual, and at that time, we adopted the penicillin breakpoints (susceptible ≥29 mm) for interpreting cefazolin zone of inhibition.

Methodological quality control was routinely performed to ensure the precision and potency of the antibiotic disc used against the standard American Type Culture Collection (ATCC) bacterial isolates *P. aeruginosa* ATCC 27853 and *E. coli* ATCC 25922.

**3. Spearman's rank correlation coefficient ($R_s$) interpretation:**

$R_s$: ±0.00-0.19 = very weak correlation; ±0.20-0.39 = weak correlation; ±0.40-0.69 = moderate correlation; ±0.70-0.89 = strong correlation; and ±0.90-1.0 = very strong correlation.
